# Proteomic profiling of CSF reveals stage-specific changes in Amyotrophic lateral sclerosis patients

**DOI:** 10.64898/2026.04.13.26350753

**Authors:** Neža Cankar, Filippa L. Qvist, Anne S. Frahm, Katrine Pilely, Kirsten Svenstrup, Stephen Wørlich Pedersen, Anne-Lene Kjældgaard, Peter Garred, Niels Henning Skotte

**Affiliations:** Department of Drug Design and Pharmacology, University of Copenhagen, Copenhagen, Denmark; Laboratory of Molecular Medicine, Department of Clinical Immunology, Rigshospitalet, Faculty of Health Sciences, University of Copenhagen, Copenhagen, Denmark; Department of Neurology, Neuroscience Centre, Rigshospitalet, Copenhagen, Denmark; Department of Neurology, Bispebjerg Hospital, Copenhagen, Denmark; Department of Neuroanaesthesiology Neuroscience Centre, Rigshospitalet, Copenhagen, Denmark; Institute of Clinical Medicine, Faculty of Health and Medical Sciences, University of Copenhagen, Copenhagen, Denmark; NNF Center for Protein Research, Faculty of Health Sciences, University of Copenhagen, Copenhagen, Denmark

**Keywords:** ALS, biomarkers, mass spectrometry, CSF proteome, proteomics

## Abstract

Amyotrophic lateral sclerosis (ALS) is a rapidly progressing neurodegenerative disease with a heterogeneous clinical presentation, complicating early diagnosis and therapeutic monitoring. To identify disease-specific biomarkers, we performed an unbiased cerebrospinal fluid (CSF) proteomic analysis in 87 ALS patients, 89 healthy controls, and 61 neurological controls using data-independent mass spectrometry. Across all quantified proteins, 399 were significantly dysregulated in ALS, including established neurodegeneration (NEFL, NEFM, UCHL1) and neuroinflammatory (CHIT1, CHI3L1, CHI3L2) markers. Correlation and pathway analyses uncovered dysregulation of immune, synaptic, and metabolic processes, with aberrant complement activation emerging as a hallmark. Complement proteins increased progressively with declining ALS Functional Rating Scale-Revised and longer disease duration, whereas early-stage markers (CLSTN3, CHAD, RELN) indicated pre-symptomatic neuronal and synaptic disruptions. Machine learning identified a minimal five-protein CSF panel (MB, ITLN1, YWHAG, FCGR3A, PGAM1) that accurately distinguished ALS patients from healthy controls, capturing disease-specific pathophysiology beyond general neurodegeneration. Our findings define a robust ALS-specific CSF proteomic signature, reveal prognostic protein candidates across disease stages, and provide a framework for diagnostic biomarker development, enabling earlier intervention and monitoring.

## Introduction

Amyotrophic lateral sclerosis (ALS) is a devastating neurodegenerative disease characterized by degeneration of the motor neurons, resulting in progressive paralysis and ultimately leading to respiratory failure and death ^1,2^. The median survival for individuals with ALS is 2-4 years ^3^. Although ∼10% of disease cases are familial (fALS) and driven by specific inherited mutations in the C9orf72 and SOD1 genes ^1,4^, most occur sporadically (sALS), which reflects a complex interplay of genetic and environmental factors ^4,5^. The clinical course of ALS is highly heterogeneous and often mimics other neurological conditions, which poses a significant diagnostic challenge ^6^. Although drugs such as riluzole and tofersen can benefit ALS patients, they provide limited disease-slowing effects, making timely diagnosis essential to initiate treatment before irreversible neuronal damage occurs ^7–10^. Thus, development and validation of novel biomarkers and drug targets in ALS are needed for several purposes including prediction, prognosis, monitoring, and clinical treatment response.

Cerebrospinal fluid (CSF) proteomics is a key approach for studying neurological disorders and offers unbiased insights into pathological changes within the central nervous system (CNS) ^11,12^. Alterations in brain-derived proteins in CSF closely reflect disease-specific changes and often precede the onset of clinical symptoms ^13^. In ALS, neurofilaments are well-established CSF and blood biomarkers, particularly neurofilament light polypeptide (NEFL) ^14–16^. This neuron-specific protein, released upon axonal injury, is elevated in ALS and correlates with disease progression ^14^. Even though NEFL is generally higher in ALS than in other neurologic or neuromuscular conditions, it lacks disease specificity ^17^. Its levels are also elevated in several other neurodegenerative diseases (eg. Huntington’s and Alzheimer’s disease), disease mimics, as well as during healthy aging ^13,17,18^. This biomarker also shows stage- and time-dependent variation, with a recent report showing that plasma NEFL rises sharply in asymptomatic individuals prior to disease onset but gradually declines after ALS onset ^16^. Moreover, NEFL primarily reflects the extent of neuronal injury ^18^ rather than the mechanisms unique to ALS. Clinical trials of the antisense oligonucleotide tofersen demonstrated reductions of NEFL in plasma and CSF following treatment, yet these changes were not accompanied by significant improvements in ALS Functional Rating Scale-Revised (ALSFRS-R) or slower disease progression ^9^. Although increased NEFL in CSF reflects neurodegeneration, additional biomarkers that capture ALS-specific molecular mechanisms and provide greater disease specificity are needed. Beyond neurofilaments, a variety of other protein candidates have been studied for their diagnostic relevance in ALS, including markers of inflammation, oxidative stress, synaptic dysfunction and neurodegeneration ^15,19^. Immune-related markers have gained most importance as diagnostic markers, with growing evidence that supports neuroinflammation as a central process in ALS ^14,19,20^. In particular, elevated CSF levels of CHIT1, CHI3L1 and CHI3L2 indicate a significant role of neuroinflammation in ALS pathophysiology ^19–21^. Likewise, increased levels of complement proteins, which are integral components of the innate immune system, have been found in the CSF of ALS patients ^22,23^. This points to a broader involvement of the immune system in ALS pathophysiology, however the potential of immune-related biomarkers for disease monitoring remains underexplored.

To characterize the CSF proteomes of ALS patients, we employed an unbiased proteomic approach in a large, well-characterized cohort, alongside healthy and neurological controls ^22,24^. By combining differential protein expression analysis with pathway enrichment, correlation studies, and machine learning (ML), we aimed to identify novel biomarkers. Our findings confirm the presence of multiple protein markers consistently associated with ALS pathology and immune-related processes as a prominent signature in the CSF. Notably, we identified stage-specific protein changes and demonstrated a progressive increase of complement markers in patients with lower ALSFRS-R scores and longer disease duration. In addition, we defined a panel of proteins with the potential to serve as diagnostic and prognostic biomarkers for ALS and to distinguish it from other neurodegenerative diseases. This framework provides a foundation for biomarker discovery and translational development in ALS and may guide future clinical trials.

## Results

### Study design and assessment of the CSF proteome analysis

This study included participants who had been previously enrolled in the Danish ALS collaborative study with data sourced from five separate hospitals ^22,24^. The cohort consists of 87 ALS patients (ALS), 89 healthy controls (HCtr), and 61 neurological controls (NCtr). Clinical metadata and CSF samples from all participants had previously been collected and stored in the biobank (**Fig 1 and Table 1**). To investigate CSF proteomic signatures, the samples were analyzed using data-independent acquisition mass spectrometry (DIA MS). (**Fig 1A**). In total, 2109 proteins (average 1688 per sample) were quantified with high reproducibility (Pearson’s R= 0.93-0.94) and no evidence of blood contamination ^25^ (**Fig 1B and C, Fig EV1 and Fig EV2**). After data filtering and processing, 1596 proteins were retained for analysis, where ∼70% overlapped with previous ALS proteomic datasets (**Fig 1D and Table EV1**) ^20,21^. Although each study employed a different proteomic technology, this marked overlap provides a relevant validation of the reproducibility of proteomic ALS studies. Robust quality metrics (median coefficients of variation 28.1-29.9%) ^11^ and consistency of proteomic complement measurements with enzyme-linked immunosorbent assay (ELISA) in the same ALS cohort ^24^ confirmed the reliability of our dataset (**Fig 1E and Fig EV3**). Furthermore, one marker, Ficolin-3 (FCN3), was common between the studies in the same cohort ^24^ and it showed a strong correlation across the two assays in all groups (R= 0.69-0.78, p < 0.0001), supporting the consistency of results between the technologies (**Fig 1F**). Collectively, these quality assessments and comparisons with earlier studies confirm the robustness and reliability of our CSF proteomic setup.

**Figure 1:**
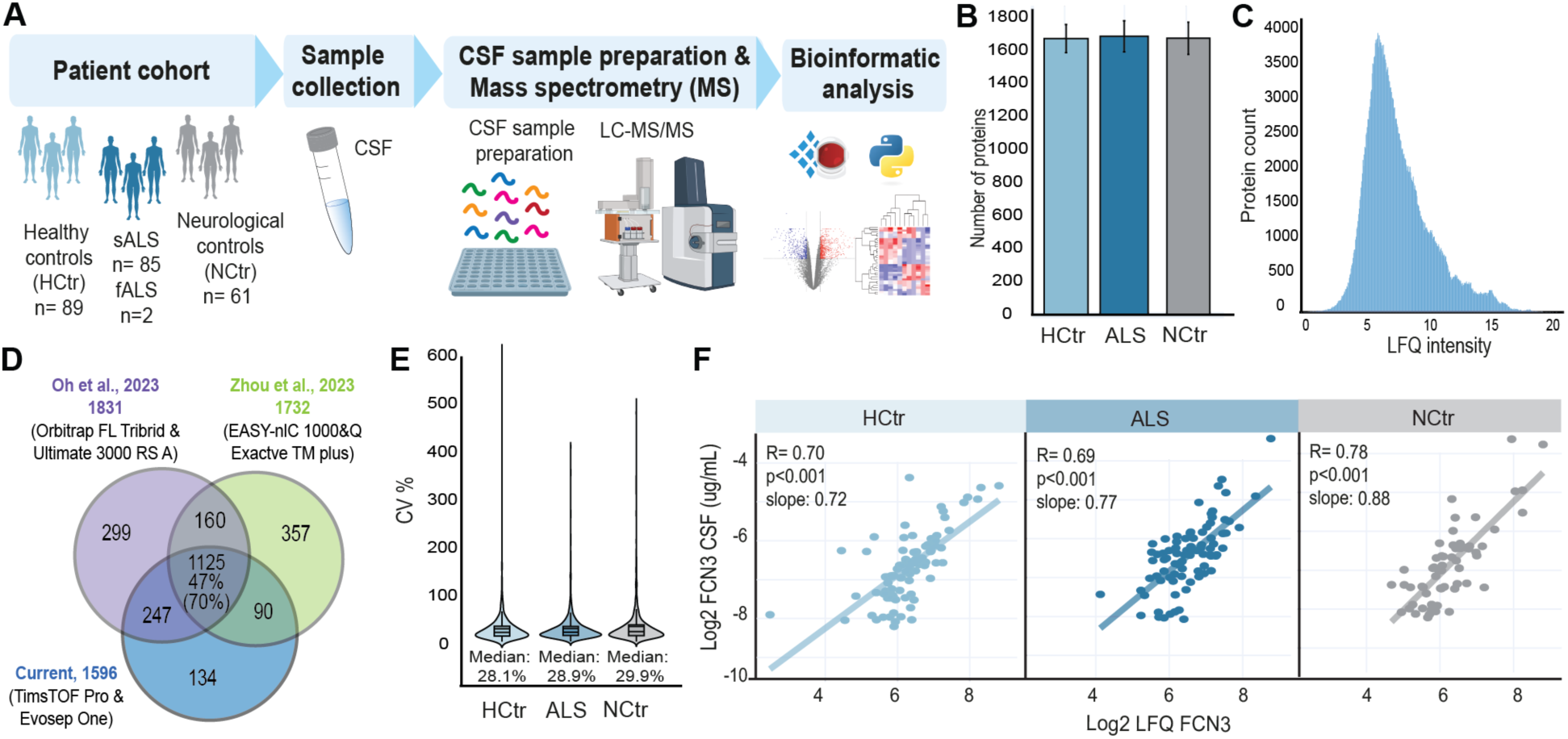
Study overview and CSF proteome characterization. A) Overview of the patient cohort and proteomic workflow; HCtr: healthy controls, NCtr: neurological controls. B) Numbers of proteins across the groups, bar plot shows ± SEM. C) Histogram plot illustrating protein distribution of processed proteomic dataset. D) Overlap between Oh et al., 2023 (purple), Zhou et al., 2023 (green) and current (blue) study, with 47% of proteins shared between all studies and 70% of our proteins detected in previous ALS datasets. E) Median coefficients of group-wise CSF sample variation in the processed dataset. F) Correlations of FCN3 protein levels in CSF across groups, as measured by mass spectrometry and ELISA assay with Pearson’s R= 0.69-0.78 and p < 0.0001 across the groups.

**Table 1:** Characteristics of participants stratified by clinical diagnosis. Values are represented as mean and standard deviation (SD). Values in brackets represent ranges of clinical scores as max an min. ALSFRS-R, revised amyotrophic lateral sclerosis functional rating scale; sALS, sporadic amyotrophic lateral sclerosis fALS, familial amyotrophic lateral sclerosis.

|  | Healthy control (HContr) | ALS | Neurological control (NCtr) |
| --- | --- | --- | --- |
| Sample number | n= 89 | n= 85 sALS, 2 fALS | n= 61 |
| Age at sampling, mean (SD) | 49.9 (11.4) | 64.4 (10.9) | 49.6 (15.5) |
| Age at symptoms onset, mean (SD) | n/a | 64.4 (10.9) | n/a |
| Bulbar onset (%) | n/a | 25 (28.7) | n/a |
| Sex (female/male) | 34/62 | 40/52 | 33/28 |
| ALSFRS-R (SD) | n/a | 35 (8.7) | n/a |
| Bulbar motor score (SD) | n/a | 9.0 (2.7) | n/a |
| Fine motor score (SD) | n/a | 8.1 (3.1) | n/a |
| Gross motor score (SD) | n/a | 8.3 (2.9) | n/a |
| Respiratory score (SD) | n/a | 9.6 (3.9) | n/a |

### Global CSF proteomes of ALS patients show significant differences in 399 unique proteins

Herein, the main objective was to identify diagnostic protein biomarkers that reflect ALS-related pathological heterogeneity within CSF. Compared to similar ALS proteomic studies, our cohort utilized a larger set of samples, allowing for a more comprehensive and well-powered analysis of the CSF proteomes ^19–21^. Consequently, we identified a higher number ^20,21^ of significantly dysregulated proteins (305 versus 97 and 33) than in two similar reports 20,21 (**Fig EV4A and Table EV1**). Additionally, an independent comparison with a proteomic study that included a cohort with a broader spectrum of ALS genetic backgrounds ^15^ revealed 212 unique proteins within our dataset (**Fig EV4B and Table EV1**), which indicates a comprehensive detection of disease-specific proteins. Consistent with this, examination of significant proteome changes across the three groups using principal component analysis (PCA) revealed a clear separation of ALS from both control groups (**Fig 2A**). To first establish a baseline proteomic characteristic in non-ALS individuals, a differential analysis between healthy and neurological control was utilized, revealing 97 protein markers (**Fig EV5 and Table EV2**). Here, a noteworthy finding was elevated levels of TREM2 in the neurological control group, a receptor that is selectively expressed by brain microglia ^26^. Given that the neurological control group exhibited some clinical features before hospitalization, but subarachnoid hemorrhage was ruled out ^22^, elevated TREM2 protein likely reflects microglial activation. This may be linked to mild immunological alterations, although the underlying cause remains to be further explored. Furthermore, the ALS group showed 346 significantly altered proteins compared to healthy controls and 147 compared to neurological controls (**Fig 2B, C and Table EV2**). 94 dysregulated proteins were shared between comparisons, and among most prominent, six proteins were strongly altered relative to both non-ALS groups, representing core markers (**Fig 2D**). This set included two neuron-enriched neurofilament proteins, NEFL and neurofilament medium polypeptide (NEFM), which have become established markers of ALS disease and are indicative of axonal damage ^14,16,17^. Another notable protein was ubiquitin carboxy-terminal hydrolase L1 (UCHL1), a multifunctional enzyme involved in the ubiquitin-proteasome system ^27^. Interestingly, CSF concentration of UCHL1 is significantly higher in ALS patients than in those with other neurodegenerative conditions ^28^. In addition, several proteins from the chitinase family (CHIT1, CHI3L1, and CHI3L2) were also found to be elevated in ALS CSF, aligning with previous proteomic findings ^19,20,29^. These neuroinflammatory markers have been increasingly associated with glial activation and innate immune responses, further supporting the contribution of neuroinflammation to ALS pathology ^19,20^. Thus, our findings recapitulate well-established protein biomarkers linked to ALS pathology that provide the basis for diagnostic framework.

**Figure 2:**
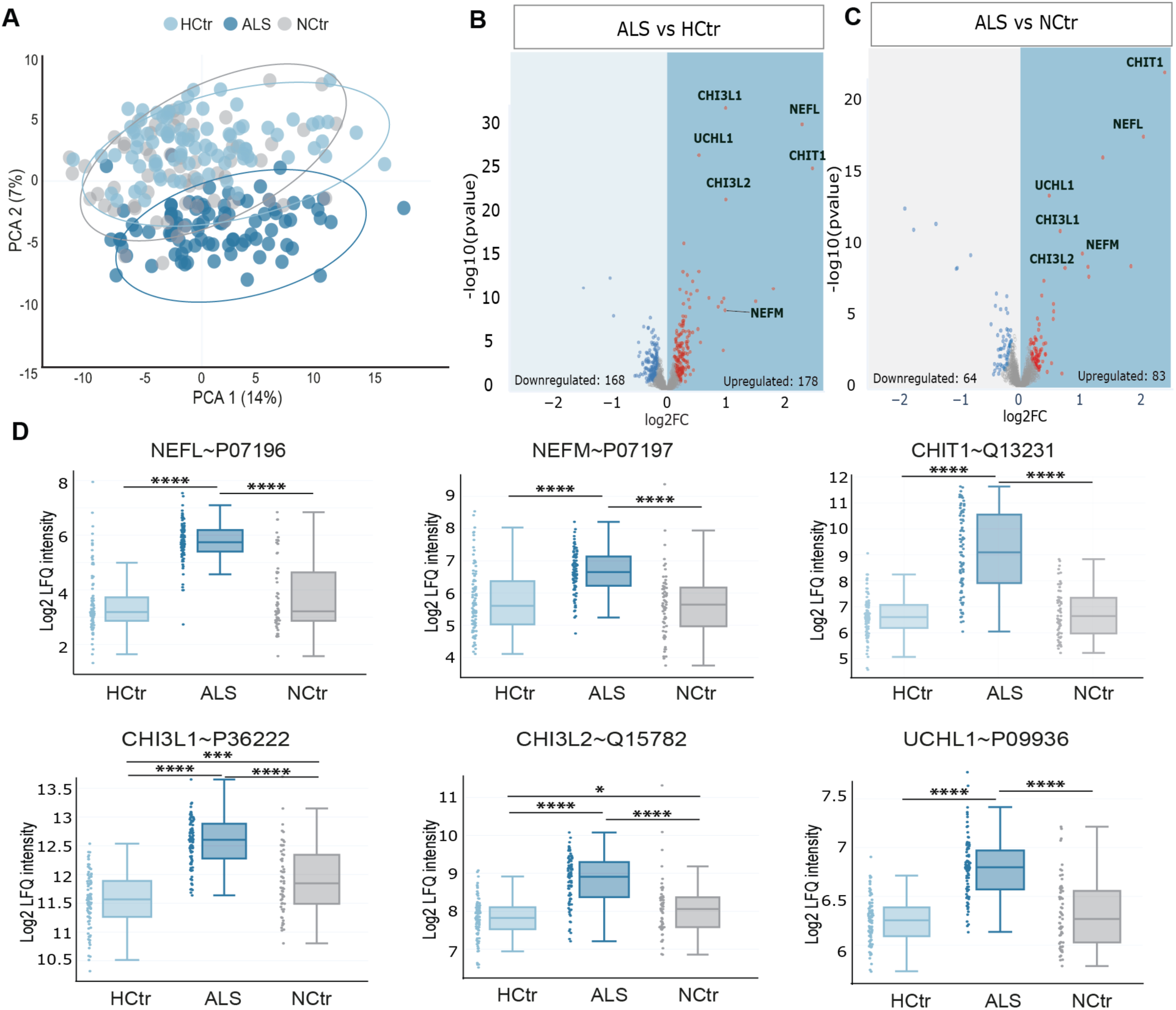
CSF proteomes from ALS patients reveal significant differences in protein regulation. A) PCA of significant proteins separates ALS patients from HCtr and NCtr. B-C) Volcano plots indicating differentially regulated proteins in ALS, compared to (B) HCtr and (C) NCtr. Data points indicating significant differences are shown in red for upregulation and blue for downregulation, based on a log2 fold change of ≥2 and a p-value < 0.05 (Student’s t-test with permutation adjustment). Gray points represent non-significant proteins. D) Total protein levels of six relevant and significant protein hits, shown as log2 label-free quantification (LFQ) values. Student’s t-test with permutation adjustment; NEFL (p****< 0.0001), NEFM (p****< 0.0001), CHIT1 (p****< 0.0001), CHI3L1 (p****< 0.0001, p***=0.0001), CHI3L2 (p****< 0.0001, p*=0.01), UCHL1 (p****< 0.0001).

### ALS-specific panel highlights dysregulation in immune, metabolic and synaptic markers

Identification of biomarkers specific to ALS remains a significant challenge, largely because the key proteins increased in ALS are also elevated in other neurodegenerative diseases ^13,17^. Resolving this limitation is essential, as specific biomarkers would enable earlier diagnosis and patient stratification, which is critical given the rapid progression and heterogeneity of ALS ^7^. While prior studies have highlighted general neurodegenerative and inflammatory markers ^19,20^, few have systematically explored CSF proteins uniquely altered in ALS. Therefore, we aimed to define proteins distinctively altered in ALS to identify disease-specific biomarker candidates. In total, 399 differentially expressed CSF proteins met the initial criteria (**Table EV2**). Among these, we observed 68 inflammatory response proteins previously linked to ALS, including two (CHIT1 and UCHL1) associated with disability progression and survival in ALS patients ^19^ (**Table EV2**). To prioritize proteins with potential diagnostic and mechanistic relevance in ALS, we next focused on three leading proteins that drove separation of clinical groups (NEFL, CHIT1 and Cannabinoid receptor 1; CNR1) (**Fig EV6**) and performed more detailed analysis. The three markers were correlated with the 399 significant to identify new proteins with a similar profile. Five proteins (IGHV5-51, Ig-like domain-containing protein, THBS2, DPEP2, and CHI3L2) demonstrated the strongest positive and five strongest negative correlation (MOG, VWC2, NPPC, CSPG5, and MT2A) to protein CHIT1 (**Fig 3A and Table EV2**). Similarly, the top proteins associated with NEFL were grouped into positively (ITGB2, CPPED1, UCHL1, GPNMB and APOB) and negatively correlated sets (ST6GAL2, TMEM132A, SCN3B, EFNB1 and CDH8) (**Fig 3B and Table EV2**). Notably, the positive co-varying pairs with CHIT1 and NEFL are involved in neuroinflammation, tissue remodeling, lipid and peptide metabolism, and calcium homeostasis which are processes well recognized in ALS pathology ^29–32^. In contrast, negatively correlated proteins are associated with oxidative stress, neuronal connectivity and oligodendrocyte structural changes, reflecting compensatory mechanisms aimed at maintaining neuronal integrity ^33^. A similar analysis was conducted for cannabinoid receptor CNR1, a protein that is markedly downregulated in ALS patients (**Fig 3C, Fig EV7 and Table EV2**). CNR1 is the most abundant G protein-coupled cannabinoid receptor in the brain and is primarily localized at the nerve terminals of both central and peripheral neurons ^34^. Among the proteins positively correlated with CNR1 (PRRT3, SCG2, GPR158, VGF, and GPR37L1), majority are linked to synaptic function, signal transduction, and neural development ^35–38^. In contrast, the negatively correlated proteins (Ig-like domain-containing protein, IGHV3OR15-7, IGLC7, IGHV3-15, and IGHV3-11) are dominated by immunoglobulin components pointing towards immune activity ^39^. Overall, these protein markers define an ALS-specific proteomic landscape of immune, metabolic and synaptic pathways related to disease mechanisms and expand the list of candidates with diagnostic potential.

**Figure 3:**
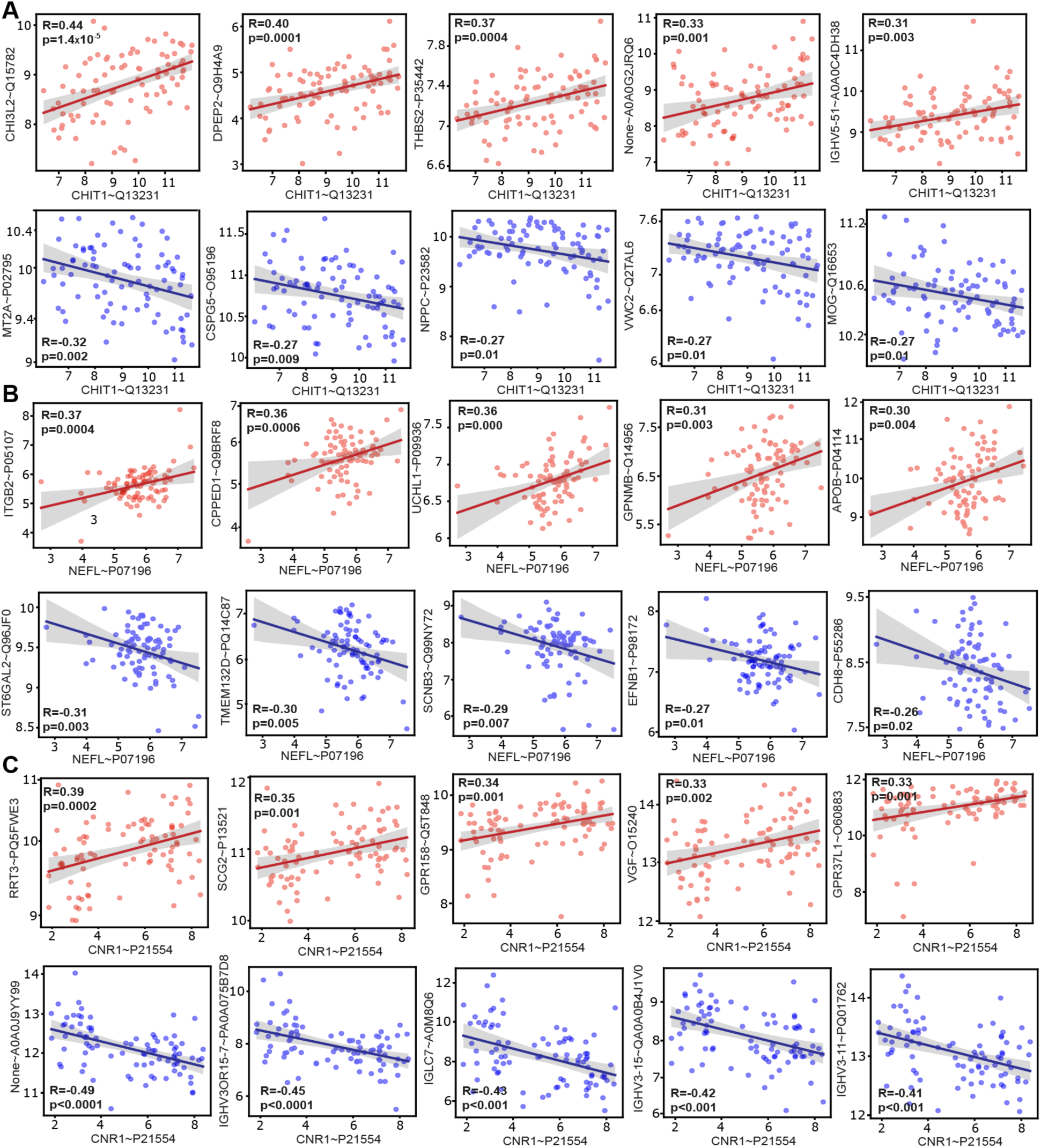
ALS-specific CSF markers show coordinated dysregulation linked to NEFL, CHIT1 and CNP1 expression. Correlation plots of A) CHIT1, B) NEFL and C) CNR1 protein levels with top positively and negatively associated ALS-specific markers, according to p-value. Protein levels are shown as log2 label-free quantification (LFQ) values. The solid lines represent linear regression fits, with shaded gray areas indicating 95% confidence intervals. Pearson correlation coefficients (R-values) are displayed within each plot.

### Aberrant complement activity is a hallmark of ALS

To further investigate the biological significance of differentially regulated proteins in this study, we went on to perform functional enrichment analyses. Establishing a baseline, a total of 428 proteins with altered CSF abundance (p < 0.05) across all groups were used to construct a global protein-protein correlation network. Here, we identified six biologically relevant clusters associated with cell-cell communication and synaptic signaling, extracellular vesicles and immune processes (**Fig EV8 and Table EV3**), underscoring that CSF composition reflects highly dynamic processes within the CNS. The three core biomarkers of ALS, proteins NEFL, CHIT1, and UCHL1 ^19,20,40^ had highly correlated expression profiles indicating that they capture similar pathological processes, such as axonal degeneration. Building on this, we performed a more detailed enrichment analysis and explored pathways that are affected by ALS. The comparison between ALS and both non-ALS groups highlighted pathways linked to mitochondrial dysfunction, platelet activity, immune responses, neutrophil degranulation, antigen binding, and lipid metabolism (**Fig 4A, Fig EV9 and Table EV3**). This agrees with previous reports that showed mitochondrial dysfunction and altered apoptotic status of platelet mitochondria in ALS patients ^41^. Moreover, dysregulated interferon gamma signaling specific to ALS group, further aligns with a recent study that found enriched interferon-associated immune signatures in human pluripotent stem cells and post-mortem spinal cord tissue ^42^. Identified neutrophil markers revealed a complex signature, suggesting a potential role in ALS pathogenesis, since they included proteins involved in migration, adhesion, and degranulation. Strikingly, a substantial proportion of upregulated pathway members (48%) were immune-related (**Fig 4B**). In combination, our findings suggest that the immune-related processes are among the main drivers of neuronal damage and disease progression in ALS.

**Figure 4:**
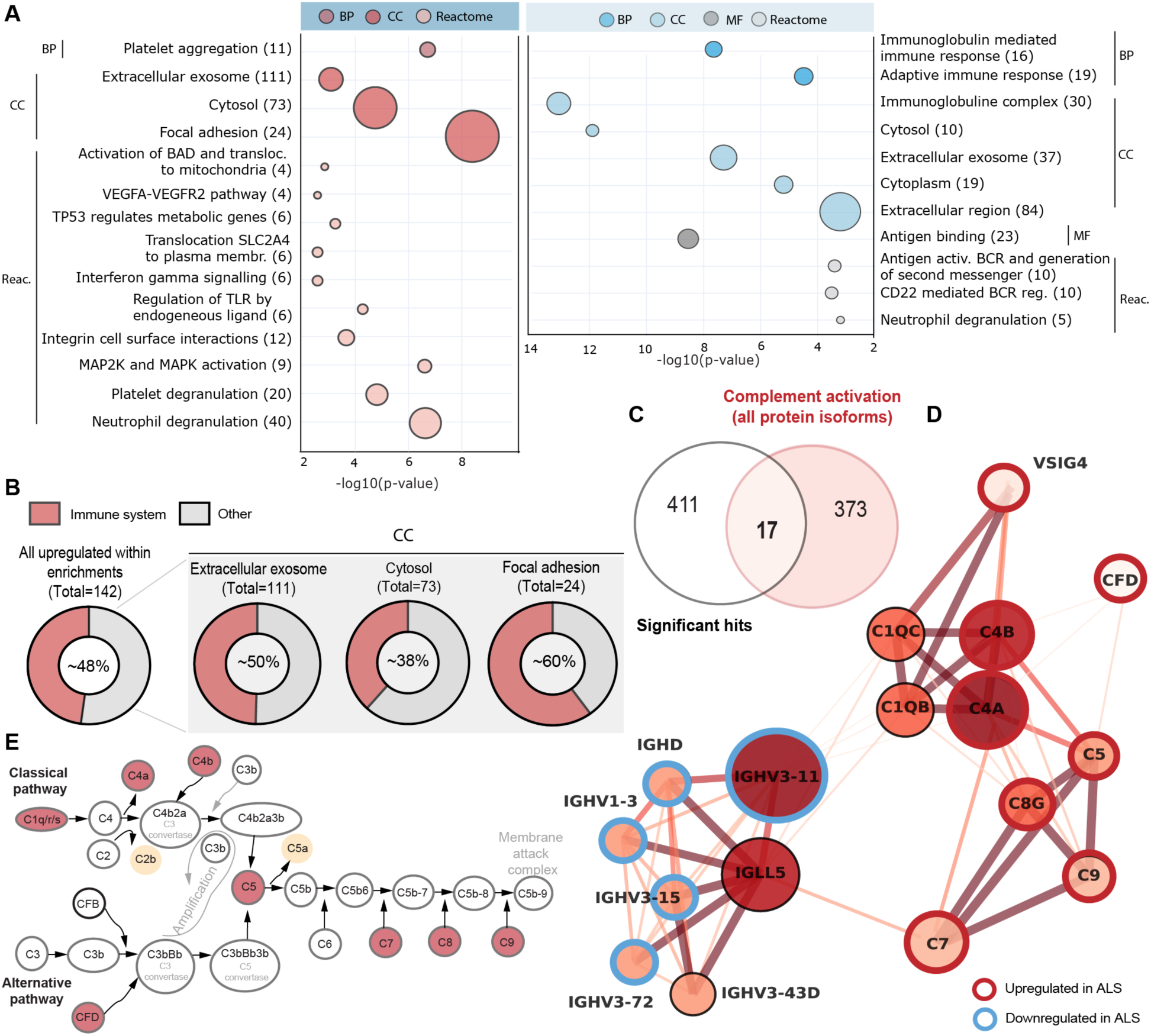
Complement protein pathway is dysregulated in the ALS group. A) Enrichment analysis of upregulated (red tones) and downregulated (blue tones) protein pathways between ALS patients and HCtr. Numbers of proteins in each pathway are presented in parentheses; BP: biological process, MF: molecular function, CC: cellular compartment. B) A donut chart displaying upregulated CC pathways in ALS group, along with the percentage of proteins associated with the immune system. Annotations were made by using the STRING online tool. C) Venn diagram of the overlap between significant hits and protein markers related to complement activation cascade, based on the UniProt database. D) Connectivity network of 17 significantly regulated proteins. Dark red tones indicate highly connected protein nodes and stronger connections between protein nodes in the network. Red circles represent significantly upregulated proteins, and blue circles indicate downregulated proteins in the ALS group. E) A scheme of classical and alternative complement pathway, with annotated significant hits from protein-protein connectivity network.

Considering the prominence of immune signatures among enriched proteins, we next examined how many of the enriched proteins were associated with the complement system (**Table EV3**), a key regulator of innate immunity implicated in ALS ^24,43^. In total, 17 proteins mediating the complement pathway were identified with close protein-protein interactions, 13 of which exhibited CSF alterations in the ALS group (**Fig 4C, D**) The first cluster comprising of immunoglobulin receptor-binding proteins was mostly downregulated and consistent with disrupted adaptive immunity, whereas the second cluster had eight upregulated proteins (C4A, C4B, C5, C7, C8G, C9, CFD and VSIG4) that are essential components of both the classical and alternative complement pathways ^44^ (**Fig 4E**). Our findings support aberrant complement function as a hallmark of ALS pathology.

### Complement proteins reflect the degree of functional impairment and disease duration

While diagnostic biomarkers are critical for early clinical diagnosis, characterizing prognostic markers is paramount for monitoring disease trajectory. To this end, we sought to uncover candidate proteins with prognostic value by examining correlations between CSF protein levels and established clinical metrics of ALS progression (ALSFRS-R, Bulbar, Fine Motor, Gross Motor, and Respiratory function) as well as with disease duration, defined as the time from symptom onset to CSF sampling. First, we focused on ALSFRS-R score as it integrates all components into a single overall measure (**Figure EV10 and EV11**). In total, 101 proteins showed correlations and the top three markers with the highest positive and negative correlation strengths include Cadherin-7 (CDH7, *p* = 1.1 × 10⁻³), Peptidyl-prolyl cis-trans isomerase (FKBP2, *p* = 2.9 × 10⁻³), Polypeptide N-acetylgalactosaminyltransferase 3 (GALNT3, *p* = 6.3 × 10⁻³), TGF-beta receptor type-2 (TGFBR2, *p* = 6.2 × 10⁻⁴), Laminin subunit beta-1 (LAMB1, *p* = 6.3 × 10⁻⁴), and Collagen alpha-1 XVIII chain (COL18A1, *p* = 1.8 × 10⁻³) (**Fig 5A and Table EV4**). Secondly, 58 proteins correlated significantly with disease duration and among these, the three with highest positive and negative correlation strengths were EGF-containing fibulin-like extracellular matrix protein 1 (EFEMP1, *p* = 1.3 × 10⁻⁴), Leucine-rich repeats and immunoglobulin-like domains protein 1 (LRIG1, *p* = 7.8 × 10⁻⁴), Complement C4-B (C4B, *p* = 1.5 × 10⁻³), CDH7 (*p* = 3.4 × 10⁻³), FKBP2 (*p* = 2.9 × 10⁻³) and Cytokine-like protein 1 (CYTL1, *p* = 2.8 × 10⁻³) (**Fig 5B and Table EV4**). Interestingly, NEFL was not among the proteins significantly correlated with either ALSFRS-R score or disease duration (**Fig EV12**), mirroring observations in tofersen studies, in which NEFL levels changed independently of ALSFRS-R trajectory ^9,45^. Together, we link specific CSF proteins with functional impairment and disease duration in ALS patients while revealing that NEFL, a key diagnostic marker, does not serve as a reliable prognostic marker in our cohort.

**Figure 5:**
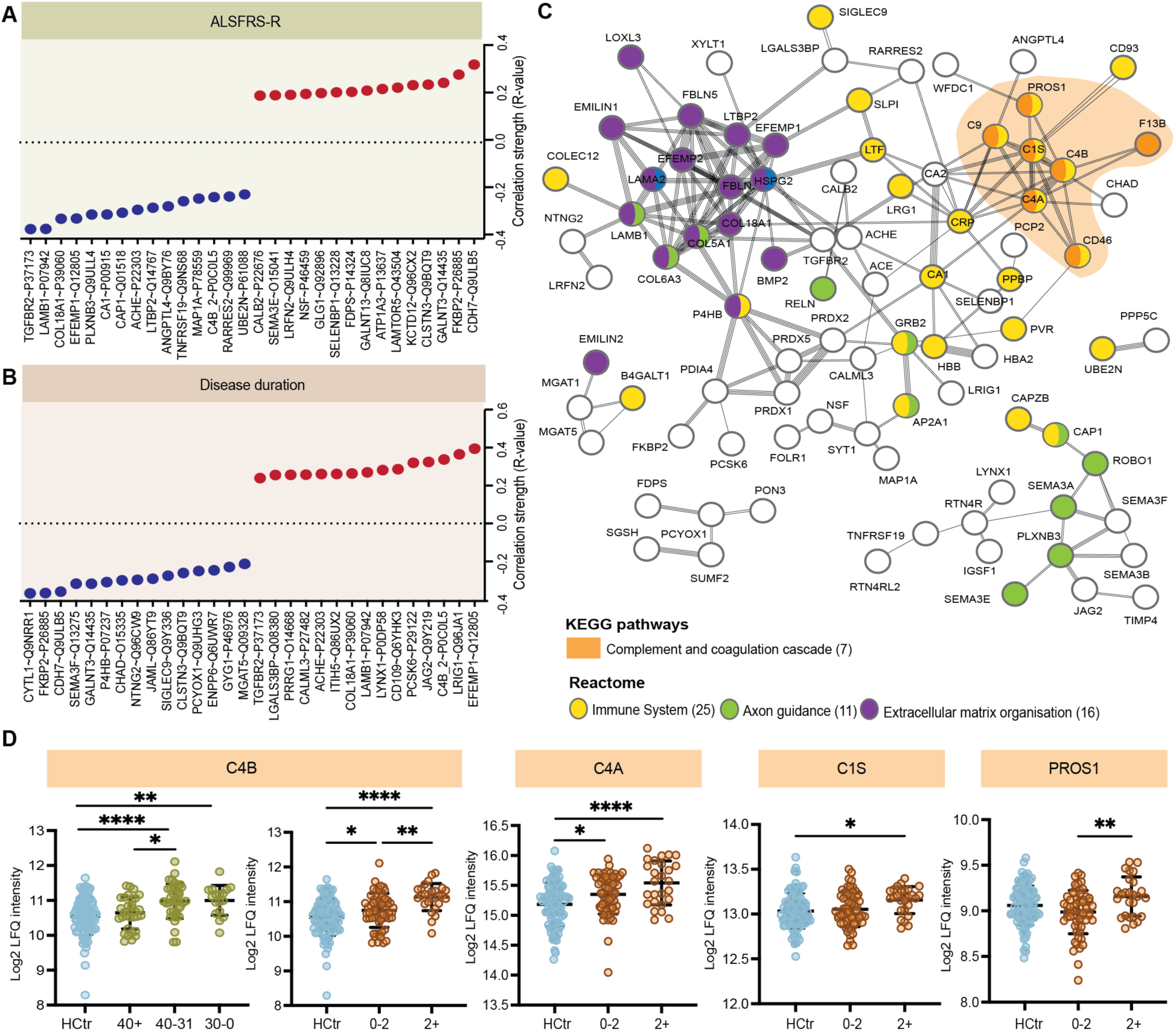
Disease duration and ALSFRS-R scores are associated with complement pathway protein expression. A-B) A lollipop plot depicting the strength and direction of correlation between CS protein levels and A) ALSFRS-R or B) disease duration in the ALS group. Top 15 proteins sorted by positive or negative correlation strength are shown. C) STRING protein interaction network of all significantly correlated proteins to ALSFRS-R and disease duration. The network illustrates 5 protein clusters, including nodes with connections. The Reactome and KEEG pathways of interest and their members are color coded. D) The proteins from the KEGG “Complement and Coagulation Cascades” pathway. Statistical comparison was performed using one-way-ANOVA with Tukey’s multiple comparison test. Total protein levels of proteins, shown as log2 label-free quantification for C4B (ALSFRS-R: p*= 0.03, p**= 0.005, p****< 0.0001 and disease duration: p*= 0.04, p**= 0.004, p****< 0.0001), C4A (p*=0.01, p****< 0.0001), C1S (p*=0.01) and PROS1 (p**=0.004). Plots show mean ± SD.

To explore the connectivity of proteins associated with ALSFRS-R and disease duration, we constructed a STRING-based protein-protein interaction network. (**Fig 5C**). Consistent with the immune-related ALS signature described above, the connectivity analysis revealed four major protein clusters, including the Complement and coagulation cascade, Immune System, Axon guidance, and Extracellular matrix (ECM) organization. Moreover, complement-associated proteins again featured prominently in the network. Next, we assessed stage-specific protein differences by stratifying ALS patients into bins based on ALSFRS-R scores (>40, 31-40 and 0-30) and disease duration (≤2 years and >2 years). These intervals were defined to reflect clinically meaningful stages of ALS, based on the non-linear decline of ALSFRS-R over time ^46^ and the typical median survival of 2 to 4 years ^1,3^. The stratification highlighted four complement-related proteins of interest (**Fig 5D**). Notably, two C4 isotypes which contribute to synapse elimination and CNS tissue remodeling ^47^, were elevated in relation to declining function and/or prolonged disease duration. Furthermore, patients with disease duration >2 years also exhibited markedly higher levels of complement cascade proteins (C4A, C1S and PROS1) compared to those with a shorter disease course (≤2 years). Collectively, we uncovered a progressive increase of complement markers in patients with lower functional scores and longer disease duration.

### Prognostic biomarker candidates in ALS

Protein changes that occur at early disease stages before substantial functional decline are of particular interest for therapeutic intervention monitoring. Using standardized ALSFRS-R scores, we investigated whether additional proteins that are independent of complement correlate with functional status and could serve as early prognostic indicators (**Table EV5**). Remarkably, Calsyntenin-3 (CLSTN3) and Chondroadherin (CHAD) proteins were already altered in patients with least affected functional scores (ALSFRS-R, >40) and shorter disease duration (≤2 years), highlighting their prognostic potential (**Fig 6A, B**). Additionally, Reelin (RELN) also distinguished ALS patients from healthy controls in early stages but did not correlate with disease duration (**Fig EV13A**). Functional characterization of these three proteins demonstrates mechanistic associations with synaptic processes. More specifically, CLSTN3 regulates axonal maintenance ^48^, CHAD is implicated in ECM remodeling of idiopathic scoliosis ^49^, and RELN is critical for regulating synaptic plasticity ^50^. Therefore, homeostatic disruptions in synaptic and extracellular processes may occur prior to comprehensive functional decline in ALS.

**Figure 6:**
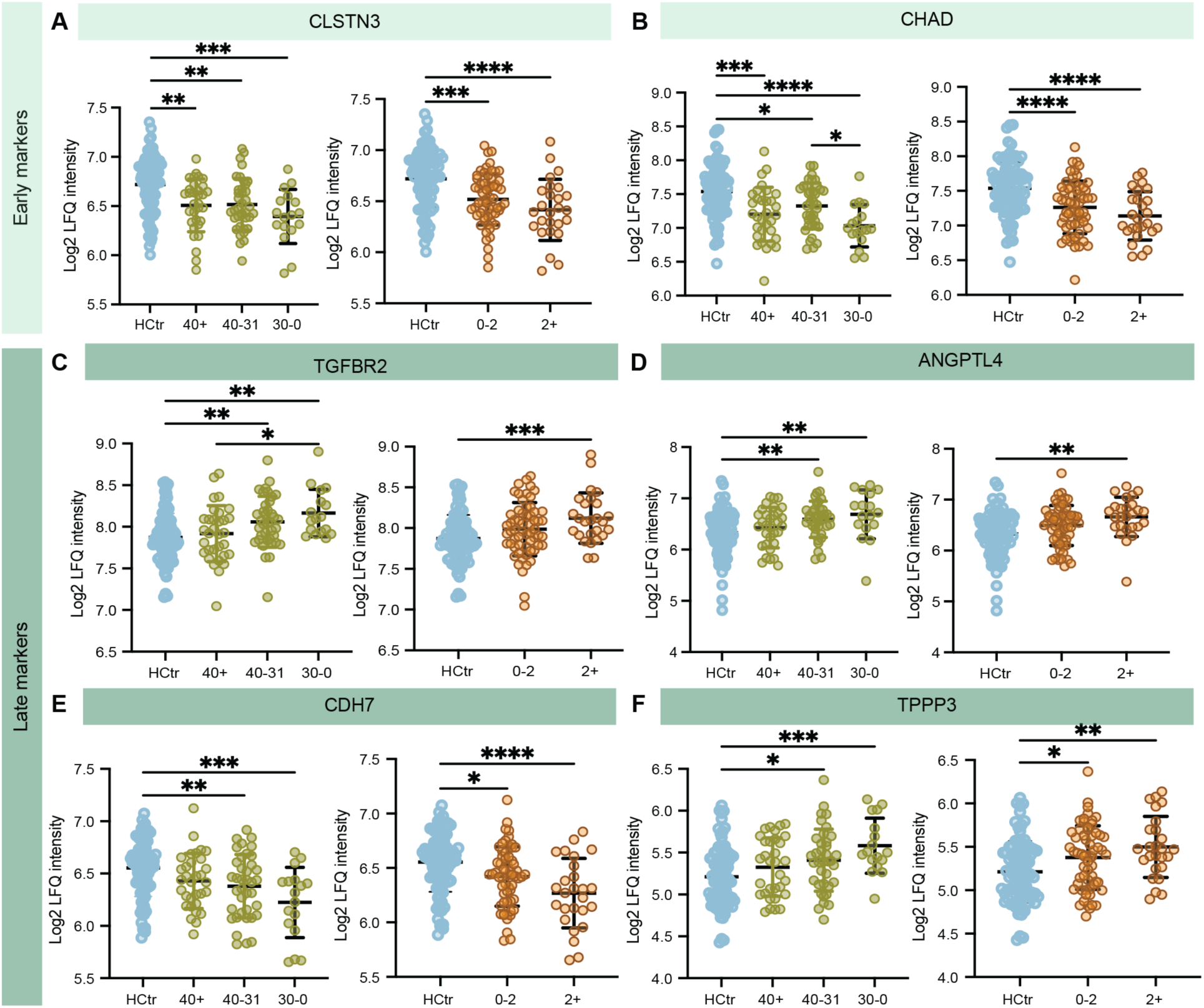
Early and late prognostic biomarker candidates in ALS. A-B) Total values for early protein markers shown as log2 label-free quantification for A) CLSTN3 (ALSFRS-R: p**= 0.001, p**= 0.001, p***=0.0001 and disease duration: p***= 0.0001, p****< 0.0001), B) CHAD (ALSFRS-R: p*= 0.03, p*= 0.01, p***= 0.0001, p****< 0.0001 and disease duration: p****< 0.0001). C-F) Total values for late protein markers shown as log2 label-free quantification for C) TGFBR2 (ALSFRS-R: p*= 0.035, p**= 0.009, p**= 0.001 and disease duration: p***= 0.0009), for D) ANGPTL4 (ALSFRS-R: p**= 0.007, p**= 0.007 and disease duration: p**= 0.001), for E) CDH7 (ALSFRS-R: p**= 0.009, p***= 0.0001 and disease duration: p*= 0.01, p****< 0.0001) and for F) TPPP3 (ALSFRS-R: p*= 0.02, p***= 0.0007 and disease duration: p*= 0.02, p**= 0.0001). Statistical comparison was performed using one-way-ANOVA with Tukey’s multiple comparison. Plots show mean ± SD.

In contrast to early-stage markers, protein changes associated with advanced functional decline may serve as late prognostic markers. The late-stage markers are especially critical in ALS because the disease progresses rapidly ^3^, and their changes show direct loss of function. Within our dataset, TGF-beta receptor type-2 (TGFBR2) showed pronounced changes in patients with lower ALSFRS-R scores (bins 31-40 and 0-30) compared to healthy controls (**Fig 6C**). Moreover, Angiopoietin-related protein 4 (ANGPTL4), Chromodomain-helicase-DNA-binding protein 7 (CHD7) and Tubulin polymerization-promoting protein family member 3 (TPPP3) were also markedly altered in the same patient groups (**Fig 6D-F**). These proteins relate to already described hallmarks of ALS, including neuroinflammation, neuronal stress responses, and vascular changes ^4,44,51^. In addition, Ubiquitin-conjugating enzyme E2 N (UBE2N) was specifically altered in patients who developed progressive clinical symptoms as reflected in lower ALSFRS-R score (**Fig EV13B)**. This change is consistent with impaired proteostasis that leads to aggregation of ubiquitinated proteins in motor neurons and represents a process unique to the final stage of the disease ^2,4^. Collectively, we identified early and late prognostic protein candidates that offer the utility for future therapeutic monitoring.

### Machine learning driven discovery of diagnostic biomarker panel in ALS

Finally, we applied ML to identify a biomarker panel with high diagnostic utility for ALS. When effective treatments for ALS become available, reducing diagnostic delays using such biomarker panels could enable earlier intervention and easier monitoring to improve patient outcomes. To establish a reference for maximal predictive performance we first evaluated efficiency of our ML model, with classification through the XGBoost algorithm ^52^, discriminating ALS patients from healthy controls based on proteins known to be associated with general neurodegeneration. For practical clinical implementation, we tested panels of different sizes to identify the smallest set of proteins that could reliably predict ALS (**Fig 7 and Fig EV14)**. Both panels with all features (Neurodegeneration-Top20) or 5 protein features (Neurodegeneration-Top5) achieved a perfect discrimination between ALS patients and healthy controls, with AUC of 1.00 on ROC curves (**Fig 7A-C and Fig EV14A-C**). The proteins with greatest influence on the class separations in both panels were NEFL, CHI3L1, CHIT1, ITLN1 and UCHL1, which validates their key relevance in ALS and demonstrates that ML can systematically pinpoint diagnostically informative proteins (**Fig 7D and Fig EV14D**).

**Figure 7:**
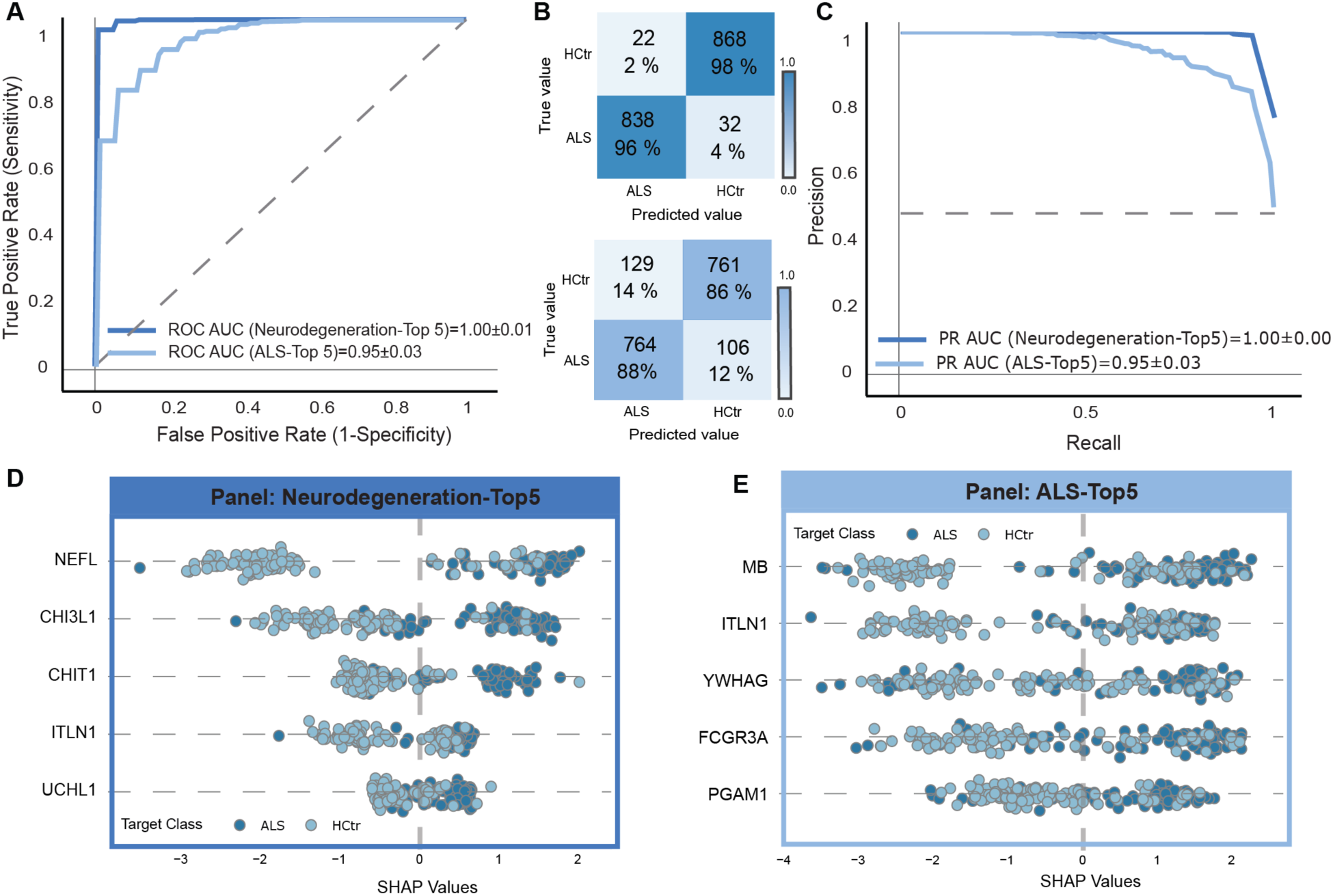
Machine learning model separates ALS from non-ALS individuals with high accuracy. A) The ROC curve was used to distinguish between ALS and healthy control subjects in two ML models, based on top 5-features. Mean ROC curves are shown in light (Neurodegeneration-Top5) and dark blue (ALS-Top5). Both models demonstrate strong performance with an area under the curve (AUC) between 1.00 - 0.95. B) Aggregated confusion matrix displaying classifier performance across all cross-validation iterations, showing correspondence between predicted and actual disease groups. Matrix illustrates ALS and healthy control model with Neurodegeneration-Top5 (above) and ALS-Top5 (below). C) The mean Precision-Recall (PR) curve for the two models is shown in light (Neurodegeneration-Top5) and dark blue (ALS-Top5). D-E) SHAP plots of top five protein features of D) Neurodegeneration model and E) ALS model. The magnitude of contribution and the likelihood for classifying ALS of each protein is indicated by the SHAP values.

Building on our initial analysis, we next repeated the ML after excluding general neurodegeneration markers (NEFL, NEFM, CHIT1, CHI3L1, CHI3L2, UCHL1) with highest differential abundance in ALS (**Fig 2**) to test whether other proteins in our dataset may provide novel predictive value. This exclusion of six proteins was also based on their shared association with axonal damage and neuroinflammatory responses which are most likely not causal but rather a consequence of the earlier pathological changes leading to neurodegeneration ^17,53^. Thus, their removal allowed us to assess whether ML could uncover ALS-specific diagnostic panels based on complementary biological processes. Remarkably, a panel with all features (ALS-Top20) achieved an AUC of 0.98, corresponding to 96% of ALS patients correctly classified (**Fig 7A-C and Fig EV14A-C**). A smaller 5-feature panel (ALS-Top5) also performed strongly, reaching an AUC of 0.95 and correctly predicting 88% of patients (**Fig 7A-C and Fig EV14A-C**). This demonstrates that accurate ML-driven classification is still possible without the most established neurodegeneration-associated ALS markers and interestingly, we identify proteins with different roles in the pathogenesis of ALS. Focusing on a minimal yet effective diagnostic panel, the ALS-Top5 predictors were MB, ITLN1, YWHAG, FCGR3A, and PGAM1 (**Fig 7E**). The muscle-derived protein MB had the greatest influence on class separations, consistent with its role in muscle metabolism and likely reflecting disease-related muscle atrophy ^54,55^. The second most important feature, Intelectin-1 (ITLN1) is an adipokine that regulates insulin-mediated glucose uptake in adipocytes and may contribute to neuroprotection. Although its role in neurodegenerative diseases is not fully understood, this protein has been shown to enhance endothelial function and revascularization after ischemia ^56^. Moreover, *in vitro* findings indicate that ITLN1 promotes the proliferation of neural stem cells and protects them from inflammation-induced damage ^57^. The remaining three protein features YWHAG, FCGR3A and PGAM1, likely indicate additional pathways affected in ALS, such as cellular signaling, inflammation and oxidative stress, all of which contribute to motor neuron degeneration ^58–60^. More specifically, changes in YWHAG point towards stress response since this protein contributes to maintaining neuronal structure under healthy condition ^61^. Its altered abundance in ALS may reflect early disturbances in intracellular signaling pathways that increase motor neuron susceptibility to degeneration ^58^. FCGR3A, an immune cell receptor, is a key mediator of antibody-driven inflammatory responses and its elevated levels likely capture the neuroinflammatory milieu characteristic of ALS ^62^. Moreover, shifts in the glycolytic enzyme PGAM1 hint at metabolic strain and oxidative pressure within CNS tissue ^59^. Together, the ALS-Top5 panel emphasizes the value of complementary biomarkers that are specific to ALS disease, whereas the Neurodegeneration-Top5 panel reflects general neurodegeneration and inflammation thus limiting the diagnostic precision. Using ML, we identified a five-protein CSF panel that accurately classified ALS status and revealed promising candidates for future multiplex diagnostic panels (**Table 2**).

**Table 2:**
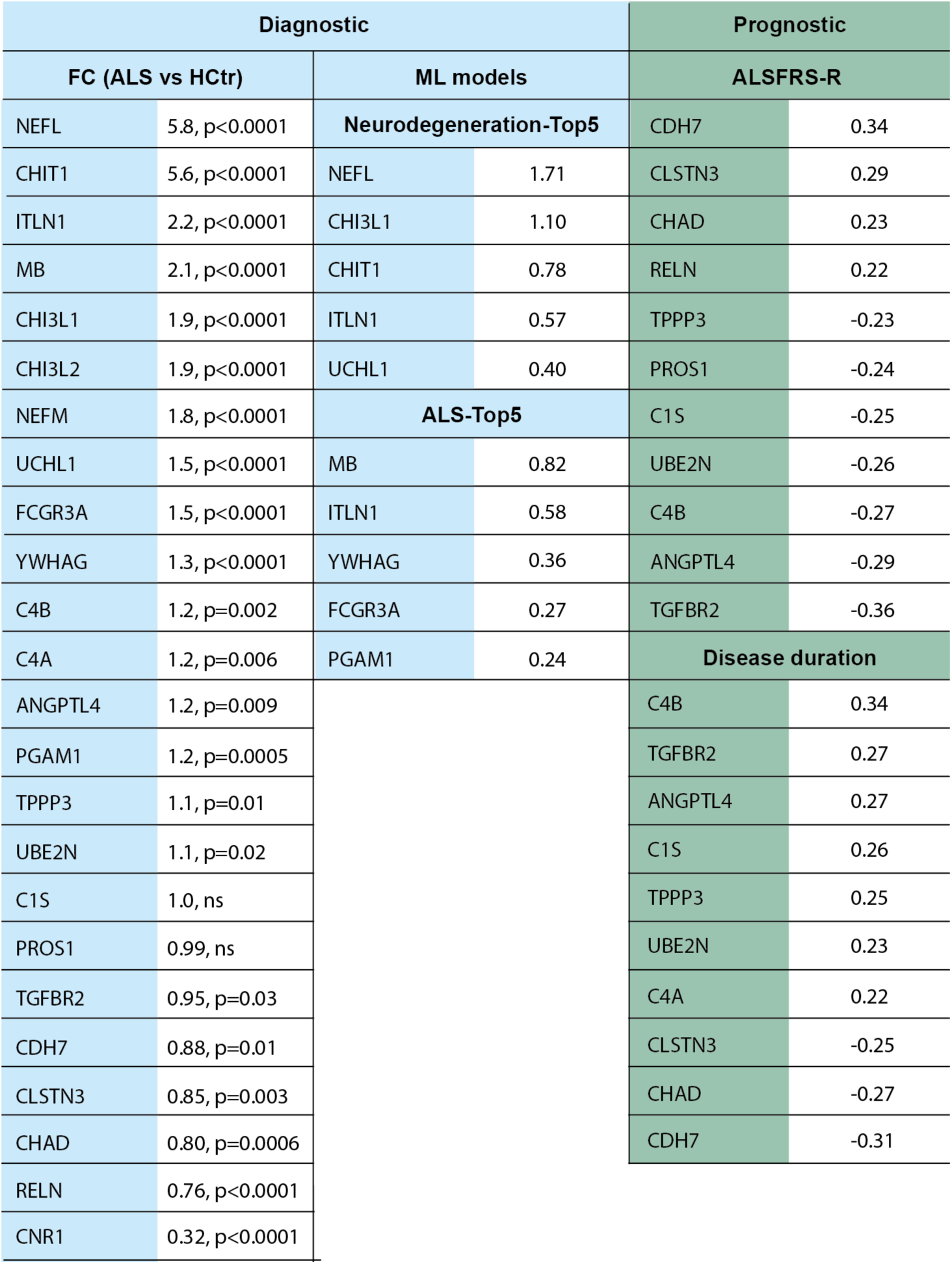
Key ALS biomarkers according to diagnostic or prognostic value. Values are shown as fold change (FC) for ALS vs HCtr together with adjusted p-values, as feature importance (Mean absolute SHAP value) for machine learning (ML) model and as a correlation strength (R-value) for both ALSFRS-R and disease duration. Prognostic markers with p < 0.05 are shown.

## Discussion

Current study provides a comprehensive characterization of the CSF proteome in a large, well-defined cohort of ALS patients compared with both healthy and neurological controls. Using DIA MS, we quantified over 2100 proteins and found 399 significantly altered in ALS, confirming known neurodegenerative markers while revealing other disease-specific signatures. Network, enrichment, and correlation analyses exhibited three mechanistic axes that define ALS CSF pathology: 1) Immune and complement dysregulation, 2) synaptic and neuronal-signaling disruption, and 3) metabolic and ECM remodeling. Complement components (C4A, C4B, C1S) showed progressive increases with declining ALSFRS-R scores and longer disease duration, whereas synaptic (CNR1, CLSTN3) and metabolic (MB, ITLN1, PGAM1) proteins reflected early functional decline and systemic reprogramming. ML analysis further shaped these patterns into an ALS-specific five-protein diagnostic panel (MB, ITLN1, YWHAG, FCGR3A, PGAM1) that accurately discriminated ALS from controls, even when canonical neurodegeneration markers were excluded. Here, our findings establish an ALS-specific CSF proteomic signature that captures stage-dependent immune activation, synaptic vulnerability, and metabolic stress, providing a mechanistic foundation for both diagnostic and prognostic biomarker development in ALS.

Our findings align with and expand upon prior ALS proteomic studies in CSF ^15,19–21^ (**Fig 1**). Consistent with earlier reports, we observed elevations of six neurodegenerative markers, including NEFL, NEFM, UCHL1, CHIT1, CHI3L1, and CHI3L2 (**Fig 2**), confirming their robustness across proteomic platforms and patient populations ^15,19–21^. Compared with earlier label-free or tandem-mass-tag-based approaches the present MS analysis leveraged a larger cohort, achieving higher proteomic depth. Our design also provided deeper coverage of differentially expressed proteins and revealed disease-stage trends that had not been identified before ^15,19–21^. Importantly, the orthogonal validation against previous datasets and ELISA immunoassay data demonstrate that these alterations are biologically reproducible rather than platform-specific artifacts ^24^. This notion is further supported by a human CSF LC-MS/MS study and an integrative multi-omics analysis across murine models and post-mortem ALS tissues, confirming our findings and the complexity of ALS biology ^23,42^. However, our dataset diverges from previous CSF reports in the magnitude and direction of specific pathway effects, showing stronger enrichment of complement activation, mitochondrial stress, and ECM remodeling (**Fig 4**). These differences likely reflect both improved statistical power and inclusion of neurological controls, which improve disease-specific contrasts. Collectively, this ALS study provides a next-generation CSF proteomic resource that bridges previous marker focused analyses with systemic insight into disease mechanisms.

While several CSF proteins are elevated in both ALS and other neurodegenerative diseases, distinguishing ALS-specific signatures is essential for diagnostic precision. For example, neurofilaments and chitinase family proteins reflect general neuronal injury and glial activation rather than ALS-unique mechanisms and are observed across multiple neurodegenerative diseases including frontotemporal lobar degeneration, Huntington’s disease and Alzheimer’s disease due to shared mechanisms between proteinopathies ^4,13,53^. To initially resolve these overlapping processes, we examined the correlation structure of the most significantly altered proteins, focusing on NEFL, CHIT1, and CNR1 as index markers for axonal degeneration, neuroinflammation, and synaptic signaling, respectively (**Fig 3**). Analysis revealed distinct co-expression modules: NEFL and CHIT1 correlated with proteins mediating immunity and lipid metabolism, whereas CNR1 was tightly linked to synaptic- and signaling-associated proteins that were selectively downregulated in ALS. These modules indicate that ALS CSF harbors distinct proteomic signatures, in line with general ALS pathology ^4,15,19,44^. Importantly, such stratification provides a biological rationale for our ML analysis, where excluding broad neurodegenerative markers enabled discovery of an ALS-specific diagnostic panel based on complementary processes. Overall, these observations reveal a disease-specific molecular architecture driving ALS pathology beyond general neurodegeneration.

Inflammation is frequently present in ALS thus representing a potential source of biomarkers for both diagnosis and tracking disease progression ^4,53^. Within this context, the complement system has emerged as a critical mediator of disease, acting both systemically and in the proximity of degenerating motor neurons ^44^. We noted dysregulation of the complement cascade as a prominent signature in the ALS CSF proteome. More specifically, functional enrichment analyses revealed that immune-related pathways made up most of the upregulated processes, with complement and coagulation components forming one of the most cohesive clusters (**Fig 4**). While several complement proteins have already been discovered in CSF of ALS patients by proteomics ^23,29,63^, we extend this list by providing new evidence of upregulated C7, C8G and VSIG4. Importantly, we identified increased levels of both classical and alternative pathway proteins including C4A, C4B, C1S, C5, C7, C8G, C9, CFD, and VSIG4 suggesting broad rather than pathway-specific alterations. These proteins are mechanistically linked to microglial and astrocytic responses, opsonization, and membrane-attack complex formation, processes known to mediate synaptic pruning and motor neuron vulnerability in ALS ^44^. Our data extend prior observations by demonstrating that complement dysregulation appears not only in the tissue but also systemically in CSF, consistent with ongoing innate immune activation in the CNS. Together, our results position aberrant complement activity as a mechanistic hallmark of ALS, linking neuroinflammation and neuronal injury at the molecular level.

Recent work demonstrated that C9orf72 is a target of autoreactive CD4+ T cells in ALS, with the balance between inflammatory and anti-inflammatory responses influencing disease progression and patient survival ^64^. Furthermore, complement elevation coupled with downregulation of immunoglobulin-binding proteins, indicates immunologic shift from adaptive to innate during ALS progression ^65,66^. We found that multiple complement cascade components including C4A, C4B, C1S, and PROS1 showed significant positive associations with disease duration and inverse correlations with ALSFRS-R (**Fig 5**). These findings support the idea of progressive complement activation as ALS advances. Stratification of patients by functional and temporal stages revealed that complement activation mirrors a gradual amplification of immune responses rather than mere persistence of inflammation. Accordingly, prior histopathological and transcriptomic findings corroborate chronic complement activation in late ALS stages ^67^. Elevated C4A and C4B may mechanistically mediate maladaptive synaptic pruning and axonal clearance, processes known to exacerbate neurodegeneration when unchecked ^44,68^. Thus, complement components offer stage-dependent biomarker candidates for monitoring disease burden and evaluating therapeutic modulation of innate immunity in ALS. In conclusion, complement activation over the disease course is reflected in dynamic changes associated with neuroinflammatory amplification and declining neuronal integrity.

Beyond immune activation, our proteomic data indicated widespread alterations in synaptic and neuronal signaling proteins, highlighting a second mechanistic axis of ALS pathology. We noted prominent downregulation of the CNR1 and its correlated partners (**Fig 3**). These included VGF, GPR158, GPR37L1, and PRRT3, all critical for synaptic homeostasis ^35–38^. CNR1 is a highly abundant metabotropic receptor in the brain and functions as a key modulator of glutamate release and excitatory neurotransmission ^34^. Its reduction in ALS CSF suggests impaired endocannabinoid signaling, which could exacerbate neuronal hyperexcitability and excitotoxicity ^1^. In animal models loss of CNR1-mediated modulation has been shown to increase neuronal vulnerability ^69,70^, whereas pharmacological or genetic enhancement of endocannabinoid signaling mitigated disease progression ^69,71^. While therapeutic potential of cannabinoids in ALS individuals remains limited, patients in a questionnaire-based survey reported improvements in both motor and non-motor symptoms ^72^. Present findings suggest that synaptic downregulation of CNR1 and other synaptic proteins co-occurs with immune activation, supporting a dual disruption of neuroimmune and neurochemical communication in ALS. We propose CNR1 as a promising therapeutic candidate for further validation, since it is directly related to a targetable disease mechanism.

In parallel with immune and synaptic changes, the ALS CSF proteome revealed alterations in energy metabolism and ECM organization, defining a third mechanistic axis of disease **(Fig 4)**. We found several metabolic proteins, including MB, ITLN1, and PGAM1, significantly elevated in the ALS group (**Table 2**). These findings emphasize changes to cellular bioenergetics, consistent with metabolomic and transcriptomic evidence of impaired energy homeostasis in ALS ^1,73,74^. In addition, proteins involved in ECM remodeling and cytoskeletal maintenance, including CHAD, TPPP3, CDH7, and RELN were also dysregulated (**Fig 6**). Because these proteins control neuronal adhesion, myelination, and cytoskeleton coupling, their alteration likely reflects progressive structural destabilization within the CNS ^4,49–51^. Such remodeling may not only compromise axonal integrity but also affect metabolism and inflammatory signaling, thereby reinforcing neurodegeneration. Together, coordinated changes in metabolic and ECM markers reveal that ALS progression involves structural reorganization beyond neurons, encompassing metabolic stress and extracellular remodeling within the CNS.

Neurofilaments and inflammatory markers provide high diagnostic value, yet they cannot fully capture the multifactorial nature of ALS ^14,75^. Higher neurofilament levels are not unique to ALS and are observed across multiple neurodegenerative conditions ^13,17^. Similarly, activation of the innate immune system represents pathogenic events in various neurodegenerative diseases ^53^, which illustrates the need for discovering disease specific protein signatures. Hence, we implemented a ML framework to evaluate the diagnostic potential of CSF proteins, comparing discriminative performance of canonical neurodegeneration markers with newly identified ALS-specific proteins (**Fig 7**). When trained with six established neurodegeneration markers, the model perfectly separated ALS patients from healthy controls. Excluding these six neurodegeneration-related features, our ML model maintained high diagnostic accuracy but captured ALS proteomic signatures beyond neurodegeneration. This is of high interest, because the resulting ALS-specific diagnostic panel comprising MB, ITLN1, YWHAG, FCGR3A, PGAM1 revealed previously underappreciated biological domains. More specifically, MB reflects skeletal muscle metabolism and denervation-associated atrophy ^4,55^. Its serum and CSF concentrations notably elevate in ALS individuals ^54^. Importantly, ALS-related muscle dysfunction results from muscular denervation and atrophy rather than fiber degeneration and necrosis, which leads to increased MB levels ^76^. Although MB does not appear to correlate with survival rate ^19,54^ and lacks specificity for distinguishing ALS from other neuromuscular conditions, it can help differentiate ALS from other neurodegenerative diseases ^54^. When combined with CSF and blood neurofilaments, MB may help refine ALS diagnosis. Furthermore, adipokine ITLN1, a regulator of glucose and lipid metabolism with pro-angiogenic effects ^77^, and PGAM1, which supports neuronal glycolysis, may capture systemic metabolic adaptation and compensatory reprogramming under oxidative stress ^59^. Such metabolic disruptions likely create an energy deficit in the skeletal muscle cells and motor neurons and drive early motor symptoms ^78^. Indeed, most patients initially present with muscle weakness, which gradually progresses to paralysis and severe disability ^4^. In parallel, YWHAG implicated in Lewy body-like hyaline inclusions in the neuronal soma, aligns with altered intracellular signaling in neurons ^58^, whereas FCGR3A indicates activation of innate immune pathways within the CNS microenvironment^60^. Overall, the exclusion of neurofilaments and inflammatory markers was critical to uncover ALS-specific features that are less likely to represent general neurodegeneration. Based on this we conclude that combining classical and novel markers could substantially enhance diagnostic specificity where distinguishing ALS from related conditions is challenging.

Identifying reliable biomarkers in ALS remains difficult due to heterogeneity of disease and the lack of clear early symptoms ^4,75^. While diagnostic markers enable earlier diagnosis of ALS, prognostic biomarkers are crucial for monitoring disease trajectory and therapeutic response ^75^. Here, a noteworthy finding was identification of proteins that show stage-specific alterations corresponding to early or late disease states. These stage-specific biomarkers not only improve disease stratification but also nominate molecular pathways that may be preferentially targeted at distinct phases of ALS progression. Early-stage protein candidates (CLSTN3, CHAD, and RELN) suggest that disruptions in synaptic and ECM homeostasis arise before extensive motor neuron loss (**Fig 6 and Fig EV13**) ^48–50^. Downregulation of CLSTN3 was already noted in ALS subjects using parallel reaction monitoring mass spectrometry method ^20,48^, yet we provide the first evidence of its prognostic relevance. Furthermore, CHAD and RELN share functional similarities with CLSTN3 since they are involved in extracellular matrix remodeling and synaptic development, thereby reinforcing a failure of synaptic functions and plasticity as a core mechanism in ALS ^33,49,79^. Moreover, several proteins (TGFBR2, ANGPTL4, CDH7, TPPP3, and UBE2N) correlated strongly with functional decline and longer disease duration, indicating their potential as late-stage biomarker candidates. Notably, TGF-β signaling pathway is increasingly recognized as a common feature of neurodegeneration. Compared to healthy controls, both TGF-β and its receptor were shown to be upregulated in postmortem tissue of ALS patients ^80^. With therapeutic evidence from non-human primates showing that targeting TGFBR2 mRNA enhances neurogenic niche activity ^81^, our findings mark TGFBR2 and TGF-β as therapeutic candidates for future interventions. In addition, we found concurrent alterations of ANGPTL4, TPPP3, and UBE2N which are indicators of late phase cellular stress and proteostasis. ANGPTL4, a hypoxia-responsive glycoprotein, may contribute to the energy imbalance and vascular dysfunction observed in ALS motor cortex and spinal cord ^82^. Experimental studies in vascular dementia rats show that overexpressing ANGPTL4 exacerbates hippocampal tissue damage through mitochondrial autophagy while its knockdown mitigates these effects ^83^. The role of TPPP3 and CDH7 proteins in ALS is less documented, but both previously been associated with neuronal integrity. TPPP3 has been implicated in axonal instability in Parkinson’s disease ^84^, while CDH7, a synaptic adhesion molecule, is downregulated in ALS post-mortem cortical synaptoneurosomes ^82^, supporting the relevance of our findings. Lastly, we observed temporal increase in UBE2N, which likely reflects compensatory activation of ubiquitin-dependent protein degradation under chronic proteostatic stress ^4,85^. In summary, early-stage ALS markers reflect synaptic and ECM dysfunction, while late-stage candidates indicate vascular, cytoskeletal, and proteostatic disturbances associated with functional decline. Together, these results reveal a temporal map of ALS progression and highlight biomarker candidates with potential for monitoring disease trajectory and therapeutic response.

Current study benefits from several methodological strengths that enhance the reliability and translational relevance of its findings. The use of a clinically well-characterized cohort with both healthy and neurological controls enabled discrimination between ALS-specific and general neurodegenerative proteomic changes. Application of deep DIA proteomics ensured broad CSF coverage and high reproducibility with robust statistical processing, multiple-testing correction, and ML models with SHAP-based interpretability, further strengthened confidence in the derived biomarker panels. Nonetheless, several limitations warrant consideration. First, our cross-sectional design limits temporal resolution and prevents direct assessment of longitudinal biomarker dynamics. Second, potential clinical confounders (e.g., medications, subclinical inflammation) and minor contributions from blood-derived proteins cannot be fully excluded despite stringent quality control. Additional investigations should replicate these findings across independent, multi-ethnic cohorts in both CSF and blood, as reliance on a single-center biobank may limit generalizability. Finally, while repeated cross-validation minimizes overfitting, external validation of the ML model remains essential before clinical implementation.

In conclusion, this work defines an ALS-specific CSF proteomic signature that captures the interplay between immune, synaptic, and metabolic dysfunction. Complement activation emerges as a central pathological axis, while ML-derived biomarker candidates reveal diagnostic patterns beyond general neurodegeneration. While some findings require further validation, we provide a mechanistic framework and translational roadmap for biomarker development in ALS, bridging discovery proteomics and clinical application.

## Methods

### Ethical approval

All the experiments were approved by Committees on Health Research Ethics in the Capital Region of Denmark (Approval number H-16017145) and Danish Data Protection Agency (File number 2012-58-0004), registered with ClinicalTrials.gov (NCT02869048) and conducted in accordance with the Declaration of Helsinki. All individuals gave written consent following receipt of verbal and written information, prior to being enrolled in the study.

### Study design

Participants within this study are above 18 years, as previously described by Kjældgaard et al. ^22,24^ and the cohort is divided into ALS patients (ALS), Neurologically healthy controls (HCtr) and Neurological controls (NCtr). ALS cohort was recruited from five Danish clinics if they met the El Escorial Revised criteria for probable or definite ALS ^86^. The HCtr group includes neurologically healthy patients who were scheduled for elective orthopedic surgery while the NCtr group consists of individuals that were initially admitted to the hospital due to suspected aneurysmal subarachnoid hemorrhage. The NCtr were only included in this study if the diagnosis of SAH was subsequently ruled out. Individuals with a history or symptoms of motor neuron disease, acute or chronic inflammatory conditions, or autoimmune disorders were excluded from both control groups (HCtr and NCtr). The CSF samples were collected as described before ^22,24^. Briefly CSF samples were immediately centrifuged at 2000 g, aliquoted and stored at −80 °C until proteomic analysis. Initial measurements of complement related proteins in plasma, serum and CSF were performed by sandwich enzyme-linked immunosorbent assays (ELISA) ^24^ while only remaining CSF aliquots were dedicated for proteomic analysis in the current study. Therefore, our cohort had 7 healthy control and 5 ALS CSF samples less than that of Kjældgaard et al ^24^. The ALS Functional Rating Scale Revised (ALSFRS-R) used to assess prognostic markers includes 12 items that assess functions commonly impacted by ALS. Each item is rated on a 0-4 scale, giving a maximum total score of 48 for individuals with normal function, with higher scores indicating better functional status.

### CSF sample preparation

CSF samples were randomized prior to sample preparation and handled in a 96-well format. 20µL of each CSF sample was processed in 30µL detergent free buffer (“E buffer”; 40mM CAA, 10mM TCEP, 100mM Tris in MS grade water) and then boiled at 95℃ for 10 minutes on a Thermomixer C (Eppendorf) at 1200 rpm. After cooling to room temperature, Trypsin and LysC enzymes were added at an order of 1:100 (w/w) based on sample protein amount, and the samples were left to digest at 37℃ and 800rpm for 2 hours. The reaction was then quenched by acidification to pH ∼2 using a 5:1 (v/v) excess of 1% TFA in isopropanol. The samples were transferred to and spun through pre-made StageTips containing 2 layers of styrenedivinylbenzenereverse phase sulfonate (“SDB-RPS”; Empore) for desalting. The trapped peptides were then washed twice with 1% TFA in isopropanol and twice with 0.2% TFA in MS grade water before being eluted into clean wells using an elution buffer of 1% ammonia and 80% acetonitrile in MS grade water. The eluted peptides were vacuum centrifuged at 45℃ in an Eppendorf concentrator plus until dry before they were resuspended in A* buffer (5% acetonitrile, 0.1% TFA in MS grade water). Concentrations were measured on a Nanodrop 2000c (Thermo Scientific), and 500ng cleaned peptides were loaded onto Evotips according to the manufacturer’s instructions.

### Mass spectrometry analysis

A human CSF-based proteome library was additionally prepared to support sample analysis. The human CSF pool used was prepared at an earlier date, but identically to the above protocol. Two human CSF pools were fractionated on an Opentrons-operated fractionation system into 24 fractions each, concatenated by 90 seconds. Samples were measured on a TimsTOF Pro (Bruker) connected to an Evosep One HPLC system by a 15cm performance column (150µm inner diameter, 1.5µM C18 beads) heated to 40℃. Samples were injected by a 44-minute (30 samples per day; ‘30SPD’) standardized gradient. The mass spectrometer was operated with positive ion polarity, a 1750V capillary voltage, 100% duty cycle and 100ms ramp time. Samples were processed by a diaPASEF method consisting of 20 isolation windows ranging from 20 to 200 m/z in width, spanning a 400 to 1200 m/z mass range as well as a 0.80 to 1.30 V·s/cm2 ion mobility range. Cycle time is estimated at 1.16s, and collision energy rose from 20eV to 59eV along with the ion mobility. The fractionated CSF pool samples were run with the same MS setup, including LC system, column and gradient, but in ddaPASEF mode. For this method, the MS1 scan range was from 100 to 1700 m/z, and ion mobility range from 0.60 to 1.60 V·s/cm2. Collision energy was set to rise from 20 to 59 eV, following the rise in ion mobility, and accumulation and ramp time were set to 100ms with 10 PASEF scans.

### Mass spectrometry data processing

Raw files from the fractionated human CSF pools were used to generate a spectral library in Spectronaut version 15.6.211220.50606, containing 23 658 precursors and 3171 protein groups. This library was used for the analysis of the DIA raw files, which were processed in Spectronaut version 17.5.230413.55965. Analyses were performed against a human FASTA file containing 20 596 entries, downloaded from Uniprot in April 2022. The search settings allowed peptides between 7 and 52 amino acids in length with Trypsin/P enzyme specificity and maximum 2 missed cleavages allowed. Methionine oxidation and cysteine carbamidomethylation were allowed as fixed modifications, and N-terminus acetyl was allowed as a variable modification. The false discovery rate (FDR) was set to 0.01 across both PSM, peptide and protein level. Protein intensities were normalized by the ‘local normalization’ option, normalizing only by precursors which were identified in all runs.

### MS bioinformatic analysis

Bioinformatic analysis including normalization, protein filtering, statistical and enrichment analysis, and visualizations was performed in Python with automated analysis pipeline ^87^. Overall, 2109 proteins were quantified, and following filtering to retain proteins present in at least 70% of group members, the dataset was used for statistical pairwise comparisons. The missing LFQ values were imputed by utilizing a mixed model with downshift and KNN (imputation_width= 0.2, standard deviation_downshift= 1.8, knn_cutoff= 0.6) ^87^. Differentially expressed proteins between the clinical groups were identified using t-test with permutation-based threshold FDR<0.05 at 250 randomizations and an s0-value of 0.01. To assess potential blood contamination, we used a list of blood protein markers as previously described ^25^ and found that the total number of detected proteins was unaffected by the levels of three common blood markers, confirming good sample quality. Sample correlations within groups were assessed with the Pearson’s correlation coefficients. Enrichment analysis on differentially expressed proteins (GOCC, GOMF, GOBP and Reactome) was performed with Fisher’s exact test and Benjamini-Hochberg FDR<0.05 cutoff. The Venn plots were illustrated with Interactive Venn online tool (https://www.interactivenn.net/), and the connectivity networks for complement specific markers and proteins correlating with both ALSFRS-R and disease duration were visualized with STRING tool (https://string-db.org/cgi/about).

### Correlation analysis

Correlation analysis of proteomic data with clinical scores (ALSFRS-R, Bulbar motor score, Fine motor score, Gross motor score, Respiratory score, Disease duration), was performed using linear regression between protein abundances (LFQ intensity) and clinical scores for each patient. Resulting metrics including slope, intercept, Pearson’s correlation coefficient (r), p-value, and standard error, were used to evaluate how specific proteins correlate with disease progression and functional impairment and to build up a list of potential prognostic markers. When correlating LFQ protein intensities with the disease duration, one participant was excluded (disease duration > 20 years).

### Machine learning model

OmicLearn (v1.4) was utilized for performing data analysis, model execution, and creation of plots and charts. Machine learning was done in Python (3.12.2). Feature tables were imported via the Pandas package (2.3.0) and manipulated using the Numpy package (2.2.0). The machine learning pipeline was employed using the scikit-learn package (1.7.0). The Plotly (6.2.0) library was used for plotting. No normalization on the data was performed. The dataset contained no missing values; hence no imputation was performed. Features were selected using a ExtraTrees (n_trees = 100) strategy with a maximum number of 20. During training, normalization and feature selection was individually performed using the data of each split. For classification, XGBoost-Classifier (version: 3.0.2, random_state = 23, learning_rate = 0.3, min_split_loss = 0, max_depth = 6, min_child_weight = 1) was used. We used a repeated (n_repeats = 10), stratified cross-validation (RepeatedStratifiedKFold, n_splits = 5) approach to classify ALS vs. HCtr. With both full data and top 5 features (all proteins included in the model) we achieved a receiver operating characteristic (ROC) with an average AUC (area under the curve) of 1.00 (0.01 std) and precision recall (PR) Curve with an average AUC of 1.00 (0.01 std). When excluding 6 proteins form the model (NEFL, CHI3L1, CHI3L2, CHIT1 and UCHL1), we achieved a robust AUC for both ROC (0.98-0.95, 0.02 std) and PR (0.95, 0.02-0.03 std) with both all proteins and top 5 features.

### SHAP analysis

To determine the influence of each feature on the successful machine learning classification, SHAP (SHapley Additive exPlanations) values were calculated for the algorithm. SHAP values are calculated for each data point in the model. A high absolute SHAP value for a feature translates to a high contribution to the model for the feature, with a positive value pointing to classifying the datapoint as ALS and a negative value as HCtr. In this way SHAP provides a robust framework for model interpretation and comparison. SHAP values were calculated using the SHAP package (0.48.0) for Python ^88^. Feature ranking was determined by the sum of the absolute SHAP value for each feature.

### Statistical analysis

All the statistical analyses except on the proteomic dataset were performed with GraphPad Prism (version 9.5.0, https://www.graphpad.com/). For statistical analysis comparing two groups with normal distribution, the unpaired t-test was used. Datasets including more than two groups were analyzed using one-way-ANOVA and Šídák’s multiple comparisons test with Tukey correction. Unless stated otherwise, the mean and SD values are reported.

## Data availability

The ALS dataset in this study is available in the following databases: a) Mass spectrometry data: ProteomeXchangeConsortium (PXD073838)

## Author contributions

NHS designed the proteomics study, supervised and guided the project, contributed to data interpretation and manuscript drafting and writing. PG provided patient samples, co-designed the study, guided the project, and revised the initial version of the manuscript. NC was responsible for data analysis and interpretation, designed and generated the figures, drafted and wrote the manuscript. FLQ performed MS sample preparation and MS-based proteomic quantification. ASF contributed to developing the ML pipeline. KP, KS, SWP, and ALK identified patients, collected ALS samples, acquired clinical data, reviewed diagnoses, critically reviewed the results, and provided critical revisions to the manuscript. All authors have read and have approved the final version of this manuscript.

## Acknowledgements

We sincerely thank all the patients who took part in this study. The authors thank (Dr. C. Delle and V. Bračič) for their critical comments on the manuscript. We thank the medical team and researcher, especially Kirsten Møller, from the ALS collaborative research group based at the Laboratory of Molecular Medicine, Department of Clinical Immunology, Diagnostic Centre, Rigshospitalet; Department of Neuroanaesthesiology, Neuroscience Centre, Rigshospitalet; Department of Neurology, Neuroscience Centre, Rigshospitalet; Department of Neurology, Bispebjerg Hospital; Department of Neurology, Roskilde University Hospital; Department of Neurology, Odense University Hospital; Department of Neurology, Aarhus Hospital; Department of Anaesthesiology, Private Hospital Gildhøj for their initiative to establish the ALS biobank. The work was supported financially by the Novo Nordisk Foundation (Grant agreements NNF14CC0001, NFF205A0063505 and NNF20SA0064201, The Jascha Foundation, Aase and Ejnar Danielsen’s Foundation, The Danish Research Council for Independent Research (DFF-6110-00489), The Danish Heart Association (15-R99-A5943-22922), The Svend Andersen Research Foundation (SARF2017), Rigshospitalet (2018-1).

## References

1. Feldman, E. L. et al. Amyotrophic lateral sclerosis. The Lancet vol. 400 1363–1380 Preprint at 10.1016/S0140-6736(22)01272-7 (2022).

2. Saberi, S., Stauffer, J. E., Schulte, D. J. & Ravits, J. Neuropathology of Amyotrophic Lateral Sclerosis and Its Variants. Neurologic Clinics vol. 33 855–876 Preprint at 10.1016/j.ncl.2015.07.012 (2015).

3. Marin, B. et al. Stratification of ALS patients’ survival: a population-based study. J Neurol 263, 100–111 (2016).

4. Masrori, P. & Van Damme, P. Amyotrophic lateral sclerosis: a clinical review. European Journal of Neurology vol. 27 1918–1929 Preprint at 10.1111/ene.14393 (2020).

5. Ingre, C., Roos, P. M., Piehl, F., Kamel, F. & Fang, F. Risk factors for amyotrophic lateral sclerosis. Clinical Epidemiology vol. 7 181–193 Preprint at 10.2147/CLEP.S37505 (2015).

6. Goyal, N. A. et al. Misdiagnosis of amyotrophic lateral sclerosis in clinical practice in Europe and the USA: a patient chart review and physician survey. Amyotroph Lateral Scler Frontotemporal Degener 25, 16–25 (2024).

7. Hinchcliffe, M. & Smith, A. Riluzole: real-world evidence supports significant extension of median survival times in patients with amyotrophic lateral sclerosis. Degener Neurol Neuromuscul Dis Volume 7, 61–70 (2017).

8. Lacomblez, L., Bensimon, G., Leigh, P. N., Guillet, P. & Meininger, V. Dose-ranging study of riluzole in amyotrophic lateral sclerosis muscle-strength deterioration in ALS patients. We have. Lancet 9013, 1425–1431 (1996).

9. Miller, T. M. et al. Trial of Antisense Oligonucleotide Tofersen for SOD1 ALS. New England Journal of Medicine 387, 1099–1110 (2022).

10. Wiesenfarth, M., et al. Effects of Tofersen Treatment in Patients with SOD1-ALS in a ‘Real-World’ Setting-a 12-Month Multicenter Cohort Study from the German Early Access Program. www.thelancet.com (2024).

11. Bader, J. M. et al. Proteome profiling in cerebrospinal fluid reveals novel biomarkers of Alzheimer’s disease. Mol Syst Biol 16, (2020).

12. Niu, L. et al. Plasma proteome profiling discovers novel proteins associated with non-alcoholic fatty liver disease. Mol Syst Biol 15, (2019).

13. Rodrigues, F. B. et al. Mutant huntingtin and neurofilament light have distinct longitudinal dynamics in Huntington’s disease. Sci Transl Med 12, (2020).

14. Kläppe, U. et al. Neurodegenerative biomarkers outperform neuroinflammatory biomarkers in amyotrophic lateral sclerosis. Amyotroph Lateral Scler Frontotemporal Degener 25, 150–161 (2024).

15. Trautwig, A. N. et al. Network analysis of the cerebrospinal fluid proteome reveals shared and unique differences between sporadic and familial forms of amyotrophic lateral sclerosis. Molecular Neurodegeneration 20, (2025).

16. Chia, R. et al. A plasma proteomics-based candidate biomarker panel predictive of amyotrophic lateral sclerosis. Nat Med 10.1038/s41591-025-03890-6 (2025) doi:10.1038/s41591-025-03890-6.

17. Gaetani, L. et al. Neurofilament light chain as a biomarker in neurological disorders. Journal of Neurology, Neurosurgery and Psychiatry vol. 90 870–881 Preprint at 10.1136/jnnp-2018-320106 (2019).

18. Meeker, K. L. et al. Cerebrospinal fluid neurofilament light chain is a marker of aging and white matter damage. Neurobiol Dis 166, (2022).

19. Dellar, E. R. et al. Data-independent acquisition proteomics of cerebrospinal fluid implicates endoplasmic reticulum and inflammatory mechanisms in amyotrophic lateral sclerosis. J Neurochem 10.1111/jnc.16030 (2023) doi:10.1111/jnc.16030.

20. Oh, S., Jang, Y. & Na, C. H. Discovery of Biomarkers for Amyotrophic Lateral Sclerosis from Human Cerebrospinal Fluid Using Mass-Spectrometry-Based Proteomics. Biomedicines 11, (2023).

21. Zhou, J. et al. Biomarkers in cerebrospinal fluid for amyotrophic lateral sclerosis phenotypes. Ann Clin Transl Neurol 10, 1467–1480 (2023).

22. Kjældgaard, A. L. et al. Amyotrophic lateral sclerosis and the innate immune system: Protocol for establishing a biobank and statistical analysis plan. BMJ Open 10, (2020).

23. Collins, M. A., An, J., Hood, B. L., Conrads, T. P. & Bowser, R. P. Label-free LC-MS/MS proteomic analysis of cerebrospinal fluid identifies protein/pathway alterations and candidate biomarkers for amyotrophic lateral sclerosis. J Proteome Res 14, 4486–4501 (2015).

24. Kjældgaard, A. L. et al. Complement profiles in patients with amyotrophic lateral sclerosis: A prospective observational cohort study. J Inflamm Res 14, 1043–1053 (2021).

25. Cankar, N. et al. Sleep deprivation leads to non-adaptive alterations in sleep microarchitecture and amyloid-β accumulation in a murine Alzheimer model. Cell Rep 43, (2024).

26. Hickman, S. E. et al. The microglial sensome revealed by direct RNA sequencing. Nat Neurosci 16, 1896–1905 (2013).

27. Falzone, Y. M. et al. Integrated evaluation of a panel of neurochemical biomarkers to optimize diagnosis and prognosis in amyotrophic lateral sclerosis. Eur J Neurol 29, 1930–1939 (2022).

28. Li, R., Wang, J., Xie, W., Liu, J. & Wang, C. UCHL1 from serum and CSF is a candidate biomarker for amyotrophic lateral sclerosis. Ann Clin Transl Neurol 7, 1420–1428 (2020).

29. Oeckl, P. et al. Proteomics in cerebrospinal fluid and spinal cord suggests UCHL1, MAP2 and GPNMB as biomarkers and underpins importance of transcriptional pathways in amyotrophic lateral sclerosis. Acta Neuropathol 139, 119–134 (2020).

30. Wong, J. K. et al. Apolipoprotein B-100-mediated motor neuron degeneration in sporadic amyotrophic lateral sclerosis. Brain Commun 4, (2022).

31. Zheng, Y., Gao, L., Wang, D. & Zang, D. Elevated levels of ferritin in the cerebrospinal fluid of amyotrophic lateral sclerosis patients. Acta Neurol Scand 136, 145–150 (2017).

32. Leal, S. S. & Gomes, C. M. Calcium dysregulation links ALS defective proteins and motor neuron selective vulnerability. Front Cell Neurosci 9, 1–6 (2015).

33. Gulino, R. Synaptic Dysfunction and Plasticity in Amyotrophic Lateral Sclerosis. International Journal of Molecular Sciences vol. 24 Preprint at 10.3390/ijms24054613 (2023).

34. Garcia, A. B., Soria-Gomez, E., Bellocchio, L. & Marsicano, G. Cannabinoid receptor type-1: Breaking the dogmas. F1000Res 5, (2016).

35. Bartolomucci, A., et al. The extended granin family: Structure, function, and biomedical implications. Endocrine Reviews vol. 32 755–797 Preprint at 10.1210/er.2010-0027 (2011).

36. Garg, A. et al. A novel syndrome with short stature, mandibular hypoplasia, and osteoporosis may be associated with a PRRT3 variant. J Endocr Soc 4, (2020).

37. Verpoort, B. et al. A postsynaptic GPR158-PLCXD2 complex controls spine apparatus abundance and dendritic spine maturation. Dev Cell 60, 2470–2486.e10 (2025).

38. Meyer, R. C., Giddens, M. M., Schaefer, S. A. & Hall, R. A. GPR37 and GPR37L1 are receptors for the neuroprotective and glioprotective factors prosaptide and prosaposin. Proc Natl Acad Sci U S A 110, 9529–9534 (2013).

39. Saleh, I. A. et al. Evaluation of humoral immune response in adaptive immunity in ALS patients during disease progression. J Neuroimmunol 215, 96–101 (2009).

40. Higginbotham, L., et al. Integrated Proteomics Reveals Brain-Based Cerebrospinal Fluid Biomarkers in Asymptomatic and Symptomatic Alzheimer’s Disease. Sci. Adv vol. 6 https://www.science.org (2020).

41. Shrivastava, M., Vivekanandhan, S., Pati, U., Behari, M. & Das, T. K. Mitochondrial perturbance and execution of apoptosis in platelet mitochondria of patients with amyotrophic lateral sclerosis. International Journal of Neuroscience 121, 149–158 (2011).

42. Hiew, J. Y., Lim, Y. S., Liu, H. & Ng, C. S. Integrated transcriptomic profiling reveals a STING-mediated Type II Interferon signature in SOD1-mutant amyotrophic lateral sclerosis models. Commun Biol 8, (2025).

43. Ganesalingam, J. et al. Combination of neurofilament heavy chain and complement C3 as CSF biomarkers for ALS. J Neurochem 117, 528–537 (2011).

44. Kjældgaard, A. L. et al. Amyotrophic lateral sclerosis: The complement and inflammatory hypothesis. Mol Immunol 102, 14–25 (2018).

45. Meyer, T. et al. Neurofilament light-chain response during therapy with antisense oligonucleotide tofersen in SOD1-related ALS: Treatment experience in clinical practice. Muscle Nerve 67, 515–521 (2023).

46. Thakore, N. J., Lapin, B. R. & Pioro, E. P. Trajectories of impairment in amyotrophic lateral sclerosis: Insights from the Pooled Resource Open-Access ALS Clinical Trials cohort. Muscle Nerve 57, 937–945 (2018).

47. Chen, Y., Chu, J. M. T., Chang, R. C. C. & Wong, G. T. C. The Complement System in the Central Nervous System: From Neurodevelopment to Neurodegeneration. Biomolecules vol. 12 Preprint at 10.3390/biom12020337 (2022).

48. Pettem, K. L. et al. The Specific α-Neurexin Interactor Calsyntenin-3 Promotes Excitatory and Inhibitory Synapse Development. Neuron 80, 113–128 (2013).

49. Haglund, L., Ouellet, J. & Roughley, P. Variation in Chondroadherin Abundance and Fragmentation in the Human Scoliotic Disc. Spine (Phila Pa 1976) 34, 1513–1518 (2009).

50. Tissir, F. & Goffinet, A. M. Reelin and brain development. Nat Rev Neurosci 4, 496–505 (2003).

51. Schreiber, S. et al. Brain Vascular Health in ALS Is Mediated through Motor Cortex Microvascular Integrity. Cells vol. 12 Preprint at 10.3390/cells12060957 (2023).

52. Chen, T. & Guestrin, C. XGBoost: A scalable tree boosting system. in Proceedings of the ACM SIGKDD International Conference on Knowledge Discovery and Data Mining vols 13-17-August-2016 785–794 (Association for Computing Machinery, 2016).

53. Abu-Rumeileh, S. et al. CSF biomarkers of neuroinflammation in distinct forms and subtypes of neurodegenerative dementia. Alzheimers Res Ther 2, (2020).

54. Vidovic, M. et al. Comparative analysis of neurofilaments and biomarkers of muscular damage in amyotrophic lateral sclerosis. Brain Commun 6, (2024).

55. Hailstones, D. L. & Gunning, P. W. Characterization of Human Myosin Light Chains Lsa and 3nm: Implications for Isoform Evolution and Function. MOLECULAR AND CELLULAR BIOLOGY (1990).

56. Kataoka, Y. et al. Omentin prevents myocardial ischemic injury through AMP-activated protein kinase- and akt-dependent mechanisms. J Am Coll Cardiol 63, 2722–2733 (2014).

57. Zhao, L. R. et al. Omentin-1 promotes the growth of neural stem cells via activation of Akt signaling. Mol Med Rep 11, 1859–1864 (2015).

58. Okamoto, Y. et al. Colocalization of 14-3-3 proteins with SOD1 in Lewy body-like hyaline inclusions in familial amyotrophic lateral sclerosis cases and the animal model. PLoS One 6, (2011).

59. Jung, H. Y. et al. Phosphoglycerate mutase 1 prevents neuronal death from ischemic damage by reducing neuroinflammation in the rabbit spinal cord. Int J Mol Sci 21, 1–15 (2020).

60. Edri-Brami, M. et al. Glycans in sera of amyotrophic lateral sclerosis patients and their role in killing neuronal cells. PLoS One 7, (2012).

61. Cetica, V. et al. Clinical and molecular characterization of patients with YWHAG-related epilepsy. Epilepsia 65, 1439–1450 (2024).

62. Liu, X. & Liang, Z. FCGR3A Drives Innate Immune Activation via M1 Macrophage Polarization in Pediatric IBD. J Innate Immun 1–28 (2025) doi:10.1159/000549190.

63. Reis, A. L. G. et al. Proteomic analysis of cerebrospinal fluid of amyotrophic lateral sclerosis patients in the presence of autologous bone marrow derived mesenchymal stem cells. Stem Cell Research and Therapy 15, (2024).

64. Michaelis, T. et al. Autoimmune response to C9orf72 protein in amyotrophic lateral sclerosis. Nature 10.1038/s41586-025-09588-6 (2025) doi:10.1038/s41586-025-09588-6.

65. Murdock, B. J. et al. Correlation of peripheral immunity with rapid Amyotrophic lateral sclerosis progression. JAMA Neurol 74, 1446–1454 (2017).

66. Mimic, S. et al. Immunology of amyotrophic lateral sclerosis – role of the innate and adaptive immunity. Frontiers in Neuroscience vol. 17 Preprint at 10.3389/fnins.2023.1277399 (2023).

67. Guttenplan, K. A. et al. Knockout of reactive astrocyte activating factors slows disease progression in an ALS mouse model. Nat Commun 11, (2020).

68. Phadke, R. A. et al. The schizophrenia risk gene C4 induces pathological synaptic loss by impairing AMPAR trafficking. Mol Psychiatry 30, 796–809 (2025).

69. Bilsland, L. G. et al. Increasing cannabinoid levels by pharmacological and genetic manipulation delays disease progression in SOD1 mice. The FASEB Journal 20, 1003–1005 (2006).

70. Gargano, A., Beins, E., Zimmer, A. & Bilkei-Gorzo, A. Lack of cannabinoid receptor type-1 leads to enhanced age-related neuronal loss in the locus coeruleus. Int J Mol Sci 22, 1–16 (2021).

71. Raman, C. et al. Amyotrophic lateral sclerosis delayed disease progression in mice by treatment with a cannabinoid. ALS and Other Motor Neuron Disorders 1, 33–39 (2004).

72. Lacroix, C. et al. Cannabis for the treatment of amyotrophic lateral sclerosis: What is the patients’ view? Rev Neurol (Paris) 179, 967–974 (2023).

73. Guo, K. et al. Longitudinal Metabolomics in Amyotrophic Lateral Sclerosis Implicates Impaired Lipid Metabolism. Ann Neurol 98, 19–34 (2025).

74. Caldi Gomes, L., et al. Multiomic ALS signatures highlight subclusters and sex differences suggesting the MAPK pathway as therapeutic target. Nat Commun 15, (2024).

75. Irwin, K. E., Sheth, U., Wong, P. C. & Gendron, T. F. Fluid biomarkers for amyotrophic lateral sclerosis: a review. Molecular neurodegeneration vol. 19 9 Preprint at 10.1186/s13024-023-00685-6 (2024).

76. Al-Sarraj, S. et al. Mitochondrial abnormalities and low grade inflammation are present in the skeletal muscle of a minority of patients with amyotrophic lateral sclerosis; An observational myopathology study. Acta Neuropathologica Communications vol. 2 Preprint at 10.1186/s40478-014-0165-z (2014).

77. Zhao, A. et al. Omentin-1: a newly discovered warrior against metabolic related diseases. Expert Opinion on Therapeutic Targets vol. 26 275–289 Preprint at 10.1080/14728222.2022.2037556 (2022).

78. Nelson, A. T. & Trotti, D. Altered Bioenergetics and Metabolic Homeostasis in Amyotrophic Lateral Sclerosis. Neurotherapeutics 19, 1102–1118 (2022).

79. Iafrati, J., Orejarena, M. J., Lassalle, O., Bouamrane, L. & Chavis, P. Reelin, an extracellular matrix protein linked to early onset psychiatric diseases, drives postnatal development of the prefrontal cortex via GluN2B-NMDARs and the mTOR pathway. Mol Psychiatry 19, 417–426 (2014).

80. Peters, S. et al. The TGF-β system as a potential pathogenic player in disease modulation of amyotrophic lateral sclerosis. Front Neurol 8, (2017).

81. Peters, S. et al. Reconditioning the Neurogenic Niche of Adult Non-human Primates by Antisense Oligonucleotide-Mediated Attenuation of TGFβ Signaling. Neurotherapeutics 18, 1963–1979 (2021).

82. Laszlo, Z. I. et al. Synaptic proteomics reveal distinct molecular signatures of cognitive change and C9ORF72 repeat expansion in the human ALS cortex. Acta Neuropathol Commun 10, (2022).

83. Yao, C. & Wang, J. ANGPTL4 promoted the cognitive impairment in vascular dementia via increasing integrin/p-Syk signalings induced mitochondrial autophagy and apoptosis in the hippocampus. Sci Rep 15, (2025).

84. Rao, M. & Chang, K.-C. Insights from TPPP3 and its family member proteins in neuronal diseases. Neural Regen Res 10.4103/NRR.NRR-D-25-00345 (2025) doi:10.4103/NRR.NRR-D-25-00345.

85. Yin, P. et al. Aged monkey brains reveal the role of ubiquitin-conjugating enzyme UBE2N in the synaptosomal accumulation of mutant huntingtin. Hum Mol Genet 24, 1350–1362 (2015).

86. Brooks, B. R., Miller, R. G., Swash, M. & Munsat, T. L. El Escorial revisited: Revised criteria for the diagnosis of amyotrophic lateral sclerosis. Amyotrophic Lateral Sclerosis 1, 293–299 (2000).

87. Santos, A. et al. A knowledge graph to interpret clinical proteomics data. Nat Biotechnol 40, 692–702 (2022).

88. Lundberg, S. M., Allen, P. G. & Lee, S.-I. A Unified Approach to Interpreting Model Predictions. https://github.com/slundberg/shap.

